# Whole genome sequence meta-analyses reveal common and rare genetic associations with critical COVID-19

**DOI:** 10.1101/2025.11.19.25340573

**Authors:** Athanasios Kousathanas, Konrad Rawlik, Erola Pairo-Castineira, Fiona Griffiths, Wilna Oosthuyzen, Sara Clohisey Hendry, Tomas Malinauskas, Guillaume Butler-Laporte, Prabhu Arumugam, Colin Begg, Marc Chadeau-Hyam, Georgia Chan, Graham Cooke, Sally Donovan, Greg Elgar, Tom A. Fowler, Peter Goddard, Charles Hinds, Peter Horby, Lowell Ling, Emma F. Magavern, Fiona Maleady-Crowe, Hugh Montgomery, Christopher A. Odhams, Peter J.M. Openshaw, Christine Patch, Augusto Rendon, Shahla Salehi, Richard H. Scott, Malcolm G Semple, Manu Shankar-Hari, Afshan Siddiq, Alex Stuckey, Charlotte Summers, Linda Todd, Susan Walker, Timothy Walsh, Helen Ward, Tala Zainy, GenOMICC Investigators, ISARIC4C Investigators, BQC19 Investigators, GENCOVID Investigators, DeCOI Investigators, POLCOVID Investigators, PMBB Investigators, Angie Fawkes, Lee Murphy, Andy Law, Veronique Vitart, Patrick F Chinnery, James F Wilson, Matthew A. Brown, Paul Elliott, Loukas Moutsianas, Mark J. Caulfield, J. Kenneth Baillie

## Abstract

In susceptible patients, COVID-19 causes life-threatening disease driven by immune-mediated inflammatory lung injury. We have previously shown that multiple common host genetic variants are significantly associated with susceptibility to critical Covid-19, ^1;2;3^ and in one case, we demonstrated that such variants can inform development of new, effective drug treatment^1;4^. Here we report an association analysis of whole-genome sequences (WGS) from 11,423 cases from the GenOMICC study and 60,628 controls, together with meta-analyses with available genome-wide data (Fig. 1). We identify a rare association signal at *SLC50A1*, primarily driven by a missense variant rs147850817 (1:155138217:G:T, Arg201Leu) that may interfere with transport function, and we identify four common association signals near *ARF1, ZNF462, KLF13* and *MVP* genes. Finally, we build a WGS-derived polygenic risk score (PRS) for critical Covid-19, which offers only marginal improvement in risk estimation for the general population but may provide clinically-valuable discrimination for extreme susceptibility.

## Introduction

The COVID-19 pandemic caused an estimated 18.2 million deaths between January 2020 and December 2021 alone ^5^. Effective medication has reduced mortality by approximately 50%, but the underlying molecular mechanisms of disease remain poorly understood.

**Fig. 1:**
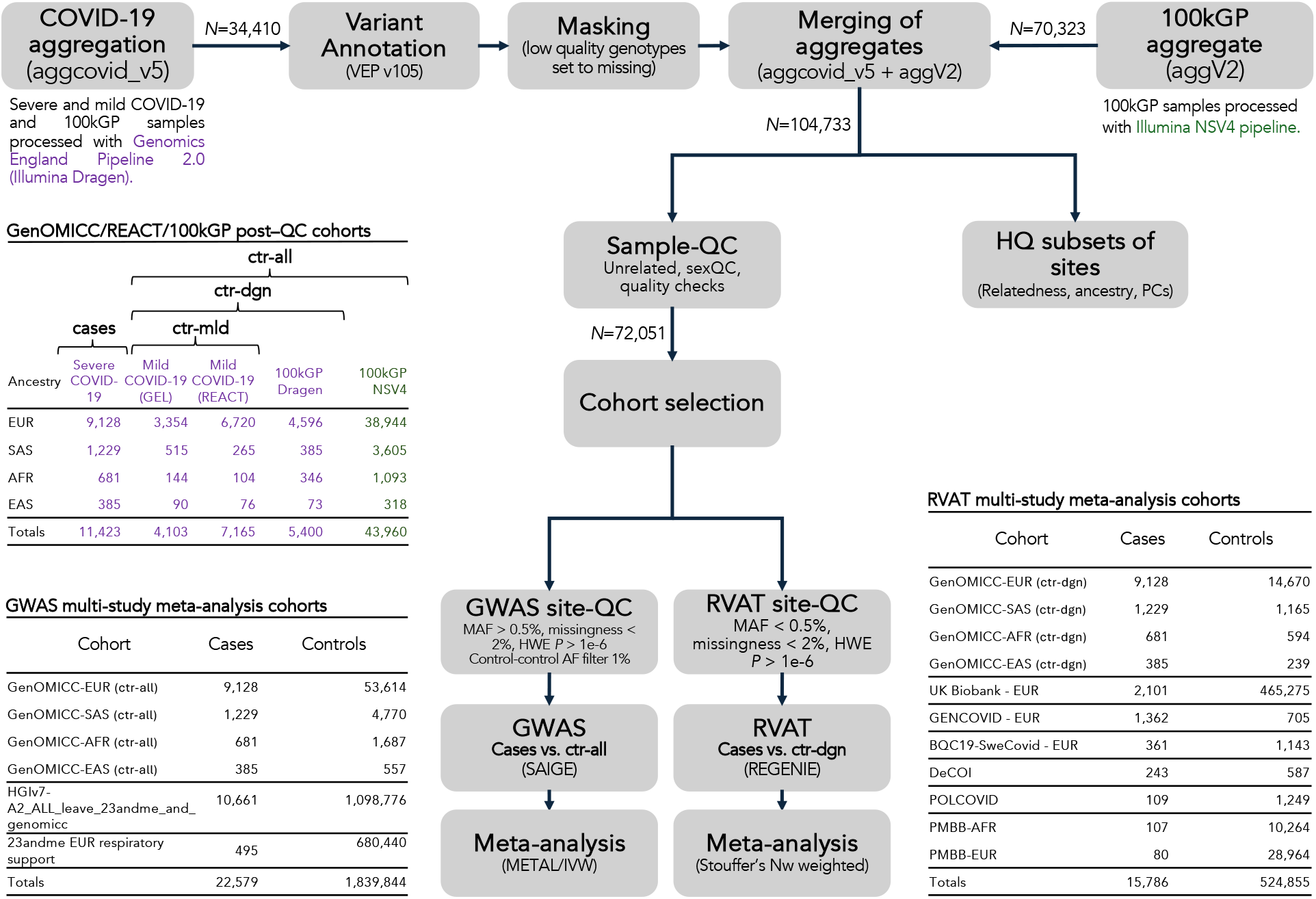
Overview of analysed cohorts and study analysis pipeline. The analysed cohorts comprised of individuals with severe COVID-19 and individuals with mild COVID-19 symptoms, along with individuals from the 100,000 Genomes Project (100kGP). COVID-19 severe and mild cohorts and a subset of 100kGP individuals were processed using the Genomics England Pipeline 2.0 (Illumina Dragen) with an additional subset of 100kGP individuals processed using a different pipeline (Illumina NSV4). Two separate aggregates were merged after masking of low quality genotypes, followed by sample quality control (sample-QC) for relatedness, sex mismatches and sample level quality. The post-quality control (post-QC) samples were divided into COVID-19 severe cases and three distinct control groups: ctrl-mld, ctrl-dgn, and ctrl-all and the sample breakdown by ancestry is shown. The ctrl-all controls set was used for GWAS analyses, while the ctrl-dgn controls set was used for rare variant aggregate testing (RVAT) analyses, as potential noise from inclusion of data from different processing pipelines is more challenging to quality control for rare variants. The ctrl-mld control set was utilized in sensitivity analyses to validate the primary findings. Additional site-wise quality control appropriate for GWAS and RVAT analyses was performed, followed by meta-analysis with other studies. A breakdown of case and control samples across studies that were included in the GWAS and RVAT meta-analyses is shown.

We and others have demonstrated that genetic factors play a critical role in determining an individual’s response to SARS-CoV-2 infection ^1;6;2;3^. Our recent large-scale meta-analysis of common variants identified 49 genome-wide significant associations associated with critical illness, highlighting the high polygenicity of severe COVID-19^3^. This observation suggests that polygenic risk scores (PRS) for severe COVID-19^7;8;9^ could be potentially useful for identifying individuals at high risk and help prioritorisation of therapeutic interventions ^7;8;9^.

Whole genome/exome sequencing (WES/WGS) enable the discovery of rare variants whose effect on disease is only detectable when aggregated, for example, across a protein-coding gene. So far, large-scale WES/WGS analyses have identified limited contributions from rare variants ^10;2^, with the most robust finding being an association between rare deleterious variants in *TLR7* ^11^.

Here we report genetic association analyses for both common and rare variants using WGS data from the largest single cohort of critical COVID-19 patients to date. We integrate these findings through meta-analysis with summary statistics from external cohort studies to identify additional common variant associations and further explore the impact of rare variants through aggregate testing. We also utilise the individual level WGS data to derive a polygenic risk score (PRS) and evaluate its performance in stratifying the general population and identifying individuals at the extremes of risk.

## Results

We conducted genome-wide association analyses (GWAS) using SAIGE^12^ for a cohort (GenOMICC) of 11,423 critically ill cases and 60,628 controls. 9,357 (82%) of the cases (7,491 (66%) with WGS data) were part of the primary analysis in our previous report ^3^. The controls included 100,000 Genomes Project (100kGP) individuals (n = 49,360) and individuals with mild COVID-19 (n = 11,268) (Methods), recruited by the GenOMICC study in partnership with the Real-time assessment of community transmission (REACT) study. A cohort breakdown by ancestry is shown in Fig. 1. We carried out separate GWAS analyses for four genetically inferred ancestry groups (African (AFR); East Asian (EAS); European (EUR); South Asian (SAS)) and using all controls (ctrl-all) for main analyses and individuals with mild COVID-19 (ctrl-mld) for sensitivity analyses (Fig. 1).

We meta-analysed GWAS results across ancestries and for each set of controls separately using inverse-variance-weighted fixed effects meta-analysis (IVW) (Methods). To maximise power for GWAS discovery, we meta-analysed the GWAS summary statistics from three studies: GenOMICC severe versus ctrl-all (this study), HGIv7-A2_ALL_leave_23andme_and_genomicc^13^, and 23andMe EUR respiratory support ^14^ (Fig. 1). To account for potential heterogeneity in variant overlap and ancestry composition across studies that could confound conditional analysis on the basis of meta-analysis summaries^15^, we defined locus associations as ±1Mbp regions surrounding top variant signals (Methods).

GWAS analysis identified 38 genome-wide significant loci (*P <* 5 × 10^*−*8^) (Fig. 2A), including putatively novel signals near *ARF1, MECOM, ZNF462, KLF13* and *MVP* genes (Table 1). We used between-study heterogeneity and independent support from meta-analysis of external studies to assess the reliability of these associations. Signals near *ARF1, ZNF462, KLF13* had support across studies and no evidence of heterogeneity (Table 1), while *MECOM* failed these tests (Table 1). The signal near *MVP*, was strongly significant in a single study (GenOMICC) but could not be confirmed because no other studies have obtained sufficient data. All novel associations had support from multiple variants in Linkage Disequilibrium (LD) (Supplementary Material), and were consistent in sensitivity analyses using alternate sets of controls (Supplementary Material). Variant-to-gene (V2G) scores from OpenTargets Genetics ^14^ identified the nearest genes as the most likely effector genes for all novel loci and were supported by cross-ancestry fine-mapping ^16^ and variant effect prediction (VEP) analysis ^17^ for credible set variants (Supplementary Material).

**Table 1:** Lead variants of novel locus association signals in the multi-study GWAS meta-analysis. Variant ids are provided as Chr:pos:ref:alt using GRCh38 coordinates with ref indicating the reference allele and alt being the effect allele. *OR, OR*_*CI*_ and *P*_*meta*_ are Odds ratios, Odds ratio 95% confidence intervals and *P*-values for the multi-study meta analysis. *P*_*het*_ is the *P*-value from heterogeneity analysis and set to “-” when there is a single contributing study (i.e. no genotype data available from other studies at this locus). *P*_*GenOMICC*_ and *P*_*hgi*_ are *P*-values for lead variants from the GenOMICC multi-ancestry meta-analysis and HGIv7-A2_ALL_leave_23andme_and_genomicc data (HGI), respectively. *P*_*GenOMICC*_ is not identical to *P* when GenOMICC was the only contributing study because of the genomic control adjustment when performing meta-analysis. Variant *3:169081067:C:T is a proxy for 3:169081115:G:GGAT with *R*^2^=0.999745 in EUR. *†* indicates the failure to validate the variant near *MECOM* in HGI at *P*_*hgi*_ *<* 0.0125. Gene column corresponds to the nearest protein-coding gene to lead variant.

| Chr:pos:ref:alt | rsid | OR | OR<sub>CI</sub> | P<sub>meta</sub> | P<sub>het</sub> | P<sub>GenOMICC</sub> | P<sub>HGI</sub> | Nearest gene |
| --- | --- | --- | --- | --- | --- | --- | --- | --- |
| 1:228089434:C:T | rs3738681 | 0.88 | 0.843-0.92 | $9.30 \times 10^{-9}$ | 0.255 | $1.00 \times 10^{-5}$ | $1.10 \times 10^{-3}$ | *ARF1* |
| 3:169081115:G:GGAT | rs112039646 | 1.15 | 1.1-1.2 | $4.81 \times 10^{-9}$ | - | $4.58 \times 10^{-9}$ | - | *MECOM* |
| *3:169081067:C:T | rs9784308 | 1.1 | 1.06-1.14 | $2.94 \times 10^{-8}$ | 0.002 | $4.53 \times 10^{-9}$ | $2.1 \times 10^{-2}\dagger$ | *MECOM* |
| 9:106709885:G:A | rs60568503 | 1.07 | 1.05-1.1 | $1.28 \times 10^{-8}$ | 0.697 | $4.65 \times 10^{-6}$ | $4.61 \times 10^{-4}$ | *ZNF462* |
| 15:31319838:G:T | rs11636034 | 0.92 | 0.9-0.948 | $1.30 \times 10^{-9}$ | 0.124 | $3.68 \times 10^{-8}$ | $2.47 \times 10^{-3}$ | *KLF13* |
| 16:29843264:G:A | rs138640006 | 2.22 | 1.67-2.94 | $2.66 \times 10^{-8}$ | - | $2.53 \times 10^{-8}$ | - | *MVP* |

**Fig. 2:**
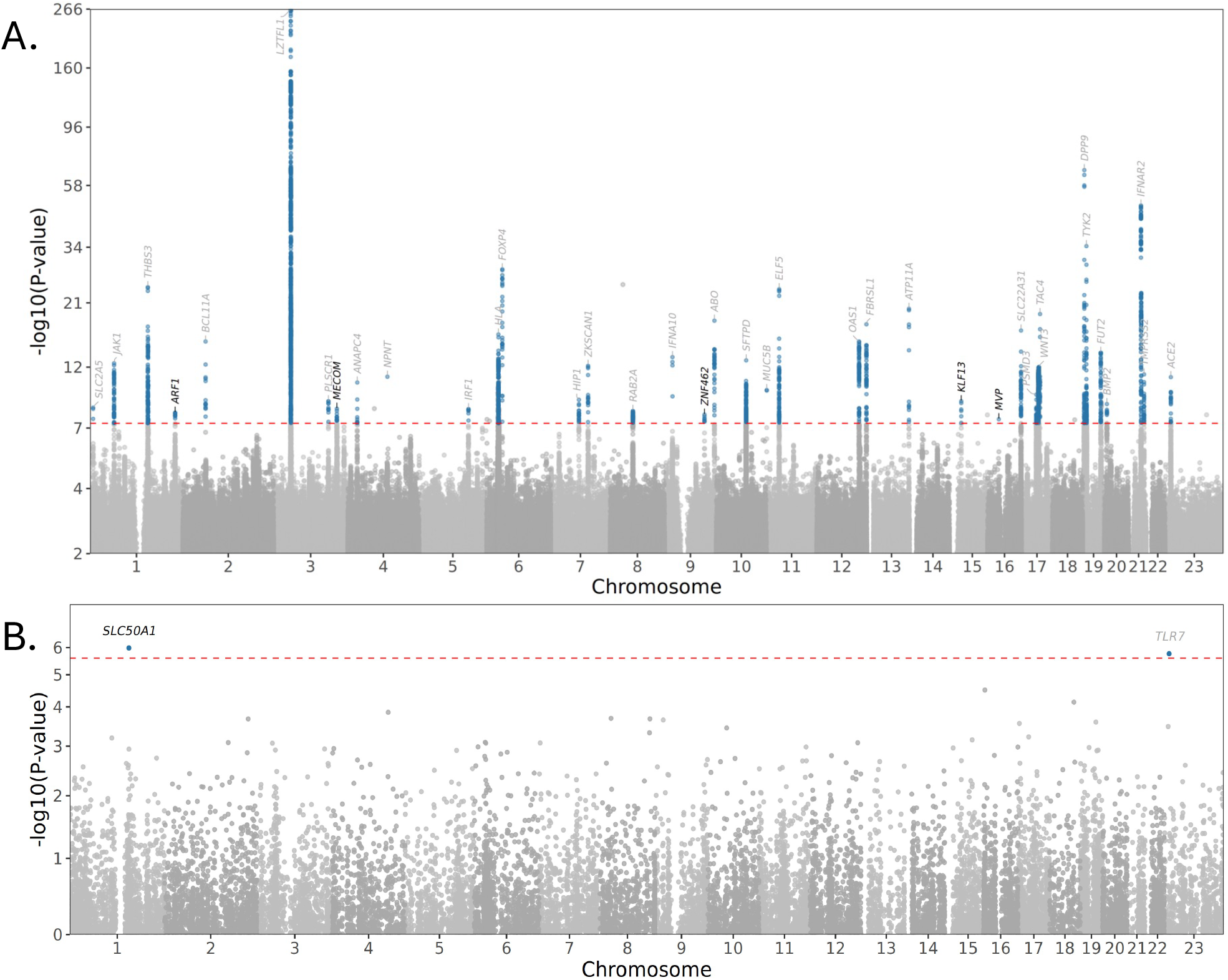
Manhattan plot of multi-study GWAS and gene-based rare variant aggregate testing (RVAT) meta-analyses. (A) GWAS results: Variants that are genome-wide significant at *P <* 5 × 10^*−*8^ (red dashed line) and within ±1Mbp of lead variants are highlighted with blue. Annotations indicate nearest gene to lead variant of each signal and text is coloured grey when previously reported and black when novel. Genome wide significant signals without support from the severe vs. mild GWAS (*P <* 5 × 10^*−*2^ and consistent effect direction) are neither highlighted nor annotated. (B) RVAT results: Genes that are gene-wide significant at Bonferroni corrected threshold *P <* 2.58 × 10^*−*6^ (red dashed line) are highlighted with blue. Annotations indicate gene and are coloured grey when previously reported and black when novel. ACAT-O gene-based *P*-values for *mild lof* mask are shown.

To assess the role of rare variants in critical illness, we conducted gene-based aggregate variant testing (RVAT) analysis with REGENIE ^18^. For this analysis, we used a subset of 28,091 individuals from our cohorts that were processed via the same alignment and variant calling pipelines, consisting of 11,423 individuals with critical COVID-19 from GenOMICC and 16,668 control individuals. The control group included 11,268 individuals from the mild COVID-19 cohort and 5,400 from the 100kGP cohort (a control set referred to as ctrl-dgn; Fig. 1; Methods). RVAT analysis was performed for each of four ancestries separately and meta-analysed with six additional studies (Fig. 1 and Supplementary material).

RVAT meta-analysis identified a significant excess burden of predicted damaging variation in *TLR7* and *SLC50A1* (*P <* 2.58 × 10^*−*6^; Fig. 2B), with *SLC50A1* being a novel RVAT association. *SLC50A1* signal was strongest in the GenOMICC European ancestry (EUR) group (*P* = 1.53 × 10^*−*5^) and nominally significant (*P <* 0.05) in two additional studies and in meta-analysis excluding GenOMICC data (Supplementary Material). *SLC50A1*, also known as *SWEET1* /*RAG1AP1*, lies within the locus indexed by common variant rs41264915 (near *THBS3*), and is partially tagged by a common novel variant signal identified through fine-mapping (see conditional analyses in Supplementary Material). We had also previously reported a significant association for *SLC50A1* with critical Covid-19 using common variant gene-level testing ^3^.

The *SLC50A1* RVAT signal was primarily driven by a single missense variant, rs147850817 (1:155138217:G:T, Arg201Leu, OR[95% CI] = 2.31[1.57-3.39]), in the largest contributing cohort (EUR GenOMICC; Supplementary Material). The Arg201Leu mutation replaces a positively charged side chain with a potentially destabilizing bulky hydrophobic one in the tightly packed hydrophilic environment facing the transport channel ^19^ (Fig. 3) and may interfere with transport function. SLC50A1 is widely expressed in human cells and tissues, including nasal epithelium and large airways. ^20^ In immune cells it is thought to play a role in recruitment of RAG complex during VD(J) recombination, an essential process in generating diversity among T- and B-cell receptors.

**Fig. 3:**
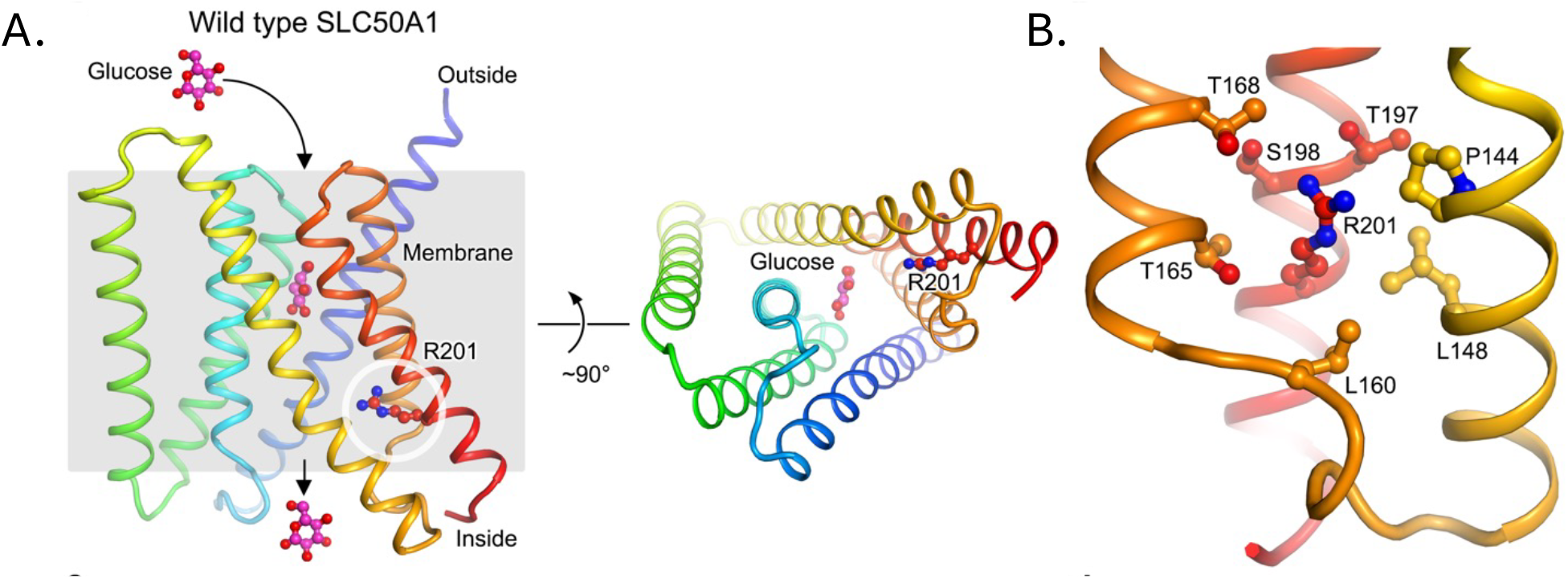
Structural analysis of the rs147850817 (Arg201Leu) mutation on SLC50A1 (A) AlphaFold model of SLC50A1 (with the N- and C-termini coloured in blue and red, respectively) with computationally docked *β*-D-glucopyranose. The side chain of Arg201 is shown as spheres and sticks. (B) zoomed-in view of Arg201 and neighbouring residues.

**Fig. 4:**
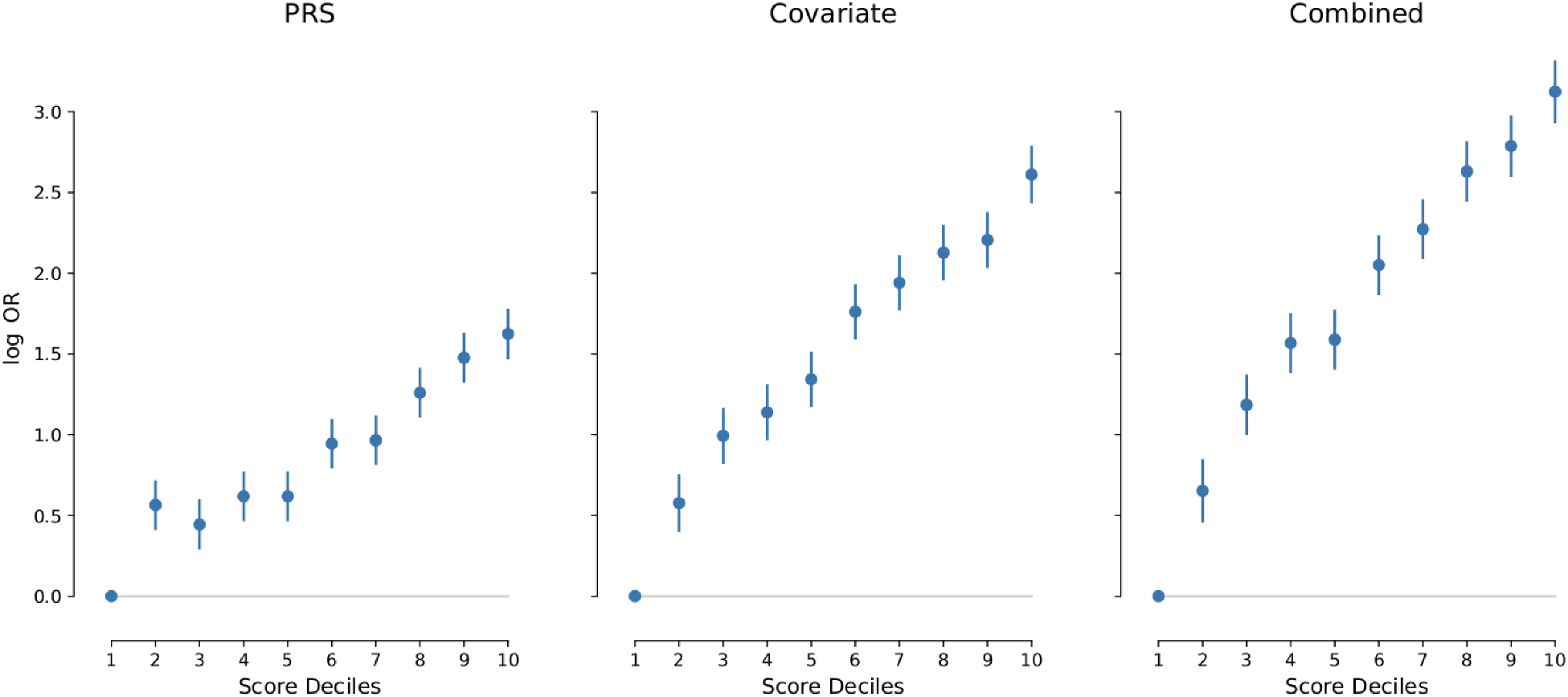
Comparison of log odds ratios of deciles of the risk score distribution relative to the bottom decile for all three considered risk scores. “PRS” indicates results from the genetic PRS score alone, “Covariate” indicates results from a risk score based on age and sex, and “Combined” indicates results from a risk score incorporating both. Odds ratios where estimated using empirical deciles in the test cohort of European ancestry.

We used results from our genome-wide association study (GWAS) on a training set comprising 80% of EUR individuals, the largest ancestry group in our study, to train a polygenic risk score (PRS) for critical covid-19. When evaluated in an independent validation set, the predictive ability of the PRS itself is limited, even within the same ancestry group (AUC 0.63, odds ratio between the bottom and top risk score decile = 2.1 (95% CI 1.03-4.16)). At a population level, PRS slightly improved risk estimation on top of a covariate based risk score (Supplementary Material), increasing validation set AUC from 0.70 to 0.73 (DeLong’s test P-value to be added).

## Discussion

In this study, we identified novel common and rare variant associations with critical COVID-19 by utilising the largest single cohort of critical COVID-19 patients with whole-genome sequencing (WGS) data to date and meta-analysis with other available datasets. We discuss the functional effects of the identified novel variants, potential effector genes, and the clinical utility of a polygenic risk score (PRS) derived from common variants.

*ARF1* is an important druggable co-factor supporting the replication of different virus families ^21;22^, including SARS-CoV2 [https://doi.org/10.1038/s41467-025-61431-8]. Inhibition of *ARF1* reduced viral replication, and knockout in mice reduced viral load and lung pathology [https://doi.org/10.1038/s41467-025-61431-8]. The lead variant rs3738681 near *ARF1* is an expression quantitative trait locus (eQTL) for both *ARF1* and *TRIM11*. Counterintuitively the T allele, identified here as protective against critical COVID-19, is associated with increased *ARF1* ^23^. Since the eQTL is based on an aggregate effect in circulating immune cells in blood, a cell-type or -state specific eQTL effect in the opposite direction may well exist, which would reverse the direction of effect. We previously observed a similar phenomenon in which an eQTL affecting PDE4A had an opposite effect in a myeloid monoculture which was not observed in bulk data, revealing a potential therapeutic target.

The same T allele is associated with decreased *TRIM11* expression in blood ^23^. *TRIM11* interferes with the replication of some viruses ^24^. Proteins encoded by both *ARF1* and *TRIM11* have been reported to down regulate interferon signalling ^25;26^. Either or both of these effects could be important: our previous genetic findings ^3^ and therapeutic trials ^27^, suggest that it is possible that interferon signalling has differential effects in early vs late disease ^28^.

We found significant disease associations for variants near the genes encoding two transcription factors: *ZNF462* and *KLF13*. The risk allele at rs60568503, near *ZNF462*, has previously been associated with with improved lung function (FEV1/FVC) ^14^. *KLF13* is a member of the Krüppel-like factor family and regulates Natural Killer (NK) cell differentiation ^29^ and can enhance the immune response to T-cell activation ^30^. The risk allele (G) at the lead variant rs11636034 of that locus is associated with increased *KLF13* expression in blood (eQTLgen); upregulated expression of *KLF13* in mice leads to proinflammatory hypercytokineaemia ^31^. The association signal at rs138640006 (*MVP*) was only detectable in patients of African ancestry. The lead variant rs138640006 is extremely rare or absent in other ancestries (Minor Allele Frequency 1.3 × 10^*−*4^ in gnomAD NFE, v4.1). Since our UK-based study has recruited by far the largest number of patients in this ancestry group, no replication set exists. *MVP* (Major Vault Protein), also known as lung disease resistance protein, is widely expressed, including in bronchial epithelium and lung tissue ^20^ and is involved in a broad range of cellular processes including including type-I interferon response and control of viral infection ^32^.

The availability of WGS data from the same sequencing and analysis pipeline provided a solid foundation for calculating a PRS for critical Covid-19. In comparison with the elements of ISARIC4C score^33^, the doubling of risk from lowest to highest decile is approximately equivalent to the impact on individual risk of having comorbid illness, or the presence or absence of signs of respiratory failure. Individuals in the top risk decile may benefit from early access to preventative measures.

Including findings near *ARF1, ZNF462*, and *KLF13* with all previously discovered associations ^3^, a total of 52 common variants have been significantly associated with critical COVID-19. A variant with an equally strong effect, near *MVP*, identified exclusively in individuals of predicted African ancestry, could not be confirmed because external data are lacking. This is a further indication that extending genetic research to include the full diversity of human populations is not only a moral, but also a scientific, imperative.

*SLC50A1* is the second gene, after *TLR7*, ^10^, in which a significant burden of aggregated rare variants is associated with critical COVID-19. Identifying the cell types and molecular mechanisms mediating the effects of these variants could provide actionable insights for therapeutic research not only in COVID-19, but also in inflammatory lung disease of other aetiologies.

## Methods

### Ethics

#### GenOMICC study

GenOMICC was approved by the following research ethics committees: Scotland ‘A’ Research Ethics Committee (15/SS/0110) and Coventry and Warwickshire Research Ethics Committee (England, Wales and Northern Ireland) (19/WM/0247). Current and previous versions of the study protocol are available at https://genomicc.org/protocol/. **Recruitment of cases (patients with COVID-19):** Patients were recruited to the GenOMICC study in 224 UK intensive care units (https://genomicc.org). All individuals had confirmed COVID-19 according to local clinical testing and were deemed, in the view of the treating clinician, to require continuous cardiorespiratory monitoring. In UK practice this kind of monitoring is undertaken in high-dependency or intensive care units. **100**,**000 Genomes project (100kGP:** The 100,000 Genomes project was approved by the East of England—Cambridge Central Research Ethics Committee (REF 20/EE/0035). Only individuals from the 100,000 Genomes project for whom WGS data were available and who consented for their data to be used for research purposes were included in the analyses. **UK Biobank study:** ethical approval for the UK Biobank was previously obtained from the North West Centre for Research Ethics Committee (11/NW/0382). The work described herein was approved by UK Biobank under application number 26041. **BQC-19:** Each participant or their legal representative (if the participant was incapable to consent) provided informed consent to the biobank. If a participant regained capacity to give consent, informed consent was obtained again directly from the participant. The study was approved by the Jewish General Hospital and Centre Hospitaler de l’Université de Montréal institutional review boards. **DeCOI:** Informed consent was obtained from each participant or the legal representative. DeCOI received ethical approval by the Ethical Review Board (ERB) of the participating hospitals/centres (Technical University Munich, Munich, Germany; Medical Faculty Bonn, Bonn, Germany; Medical Board of the Saarland, Germany; University Duisburg-Essen, Germany; Medical Faculty Duesseldorf, Duesseldorf, Germany) **Penn medicine (PMBB):** Recruitment of PMBB participants was approved under IRB protocol 813913 and supported by Perelman School of Medicine at University of Pennsylvania. **POLCOVID-Genomika (POLCOVID):** All study participants provided written informed consent and received detailed information on the study and associated risk before enrollment. The study was approved by the Bioethics Committee of the Medical University of Bialystok. **Swedish Biobank:** Informed consent was obtained for all study participants. The study was approved by the National Ethical Review Agency (Sweden) (No. 2020–01623). **23andMe study:** participants in this study were recruited from the customer base of 23andMe, a personal genetics company. All individuals included in the analyses provided informed consent and answered surveys online according to the 23andMe protocol for research in humans, which was reviewed and approved by Ethical and Independent Review Services, a private institutional review board (http://www.eandireview.com).

### Case and control cohorts

The case cohort comprises patients recruited to the GenOMICC study with confirmed COVID-19 which were deemed to require continuous cardiorespiratory monitoring by their treating clinician.

The control cohort comprises:

- A mild cohort, consisting of individuals with confirmed COVID-19 who experienced mild symptoms:
  - participants recruited as part of the Real-time assessment of community transmission (REACT) study, and
  - volunteers enrolled via a microsite who were required to self-report the details of a positive COVID-19 test, on the basis of having experienced mild (non-hospitalised) or asymptomatic COVID-19.
- Participants from the 100,000 Genomes Project (100kGP) with demographic characteristics as shown in Kousathanas et al. (2022) ^2^. For the 100kGP, we excluded individuals with haematological cancers and those who had tested positive at least once, had not tested negative on the same day, and whose test results came from hospital A&E.

For the main GWAS, we utilized the full set of controls to maximize power and employed the mild cohort for validation checks. For the gene-based rare variant analyses (RVAT), we utilized a subset of 16,668 controls (of which 11,268 were mild COVID-19 cases and 5,400 individuals were from the 100kGP) that were processed with the same alignment and variant calling pipeline as the case cohort. Cohort breakdowns are shown in Fig. 1.

### Genotype calling

For the critical and mild COVID-19 cohorts, sequencing data alignment and variant calling were conducted using Genomics England pipeline 2.0 with DRAGEN software (v.3.2.22), aligned to the GRCh38 reference genome with decoy and alternative haplotypes (ALT-aware). Genomes from the 100,000 Genomes Project cohort were processed using the Illumina NSV4 Whole Genome Sequencing Workflow (v.2.6.53.23) with the iSAAC Aligner and Starling Small Variant Caller, aligned to Homo Sapiens NCBI GRCh38 with decoys. A subset of genomes from the cancer program of the 100,000 Genomes Project was reprocessed with the COVID-19 cohort’s DRAGEN pipeline (v.3.2.22) and were included as additional controls in rare variant analyses that were more sensitive to potential batch effects than common variant analyses.

### Aggregation

We aggregated the genomic data using the Dragen gvcfgenotyper version 3.8.1 for DRAGEN processed genomes and with Illumina’s gvcfgenotyper v.2019.02.26 for Illumina NSV4 processed genomes. Variants were normalised with vt v.0.57721.

### Sample-QC

We employed the sample-QC protocol from Kousathanas et al. (2022) ^2^ by removing samples that failed four BAM-level quality control filters: freemix contamination *>* 3%, mean autosomal coverage *<* 25×, per cent mapped reads *<* 90% or per cent chimeric reads *>* 5%. We also computed additional metrics: ratio of heterozygous to homozygous genotypes, ratio of insertions to deletions, ratio of transitions to transversions, total deletions, total insertions, total heterozygous SNPs, total homozygous SNPs, total transitions and total transversions. We required that samples were within 4 median absolute deviations (MADs) of the median of each of these statistics and removed outliers. We required concordant phenotypic and genetically inferred sex, either ‘XX’ or ‘XY’ and filtered for unrelated participants (KING-robust pairwise kinship *<* 0.0442).

### Site-QC

Following Kousathanas et al. (2022) ^2^, we masked low-quality genotypes by setting them to missing using the bcftools setGT module: For autosomes, we masked genotypes having *DP <* 10 or genotype quality (*GQ*) *<* 20 or heterozygote genotypes failing an ABratio binomial test with P-value *<* 10^*−*3^. For chrX, we masked females as for autosomes. We masked male genotypes having *DP <* 5 or *GQ <* 20. After masking, we removed variant sites with missingness *>* 2% for GWAS analysis and *>* 5% for rare variant aggregate testing analysis.

### Genetic ancestry prediction

To infer the genetic ancestry of each individual we followed the same approach as in Kousathanas et al. (2022) ^2^. Briefly, we performed principal component analysis (PCA) on unrelated 1KGP3 individuals with GCTA v.1.93.1 beta using high-quality common SNPs, and inferred the fist 20 PCs. We subsequently calculated loadings for each SNP, and used these to project individuals onto the 1KGP3 PCs. We then trained a random forest algorithm from the R package randomForest with the first 10 1KGP3 PCs as features and the super-population ancestry of each individual as labels. These were ‘AFR’ for individuals of African ancestry, ‘AMR’ for individuals of American ancestry, ‘EAS’ for individuals of East Asian ancestry, ‘EUR’ for individuals of European ancestry and ‘SAS’ for individuals of South Asian ancestry. We assigned individuals to a super-population when class probability *≥* 0.8. Individuals for whom no class had probability *≥* 0.8 were not included in the analyses.

### PCA

Following Kousathanas et al. (2022) ^2^, we used sets of high quality independent sites to perform PCA using GCTA ^34^ separately for common (maf*≥*0.05) and rare (mac*≥* 5 & maf*<*0.01) variants and for each of the four population cohorts based on predicted ancestry (AFR, EAS, EUR, SAS), using unrelated individuals. We projected the inferred PCs from unrelated individuals to the entire cohorts for the first 20 common and the first 20 rare variant PCs.

### Genome-wide association study analyses (GWAS)

We performed GWAS analysis using SAIGE version 1.07^12^ for each ancestry for sites with maf *>* 0.005. We used sex, age, age^2^, age × sex, the first 20 principal components from high quality common variants.

### GWAS site-QC

We filtered variants showing evidence for differential missingness or deviations from Hardy-Weinberg equilibrium using plink version 1.9 and with *P <* 10^*−*5^ and *midP <* 10^*−*6^, respectively. For analyses using all the 100kGP controls (ctrl-all), we performed an additional control-control comparison to filter sites. 100kGP consisted of individuals processed with two different pipelines, the NSV4 Illumina pipeline and the Genomics England pipeline 2.0. We compared genomic data for a subset of these for which we had sequenced with both platforms and retained variants with relative allele frequency difference between platforms *<* 1%.

### Multi-ancestry and multi-study meta-analyses

We performed an inverse-variance weighted meta-analysis across all ancestries in our cohort (GenOMICC severe vs ctrl-all) using METAL software ^35^. Then, we meta-analysed the results of GenOMICC with HGIv7 critical (A2) leave-GenOMICC-out summary statistics (REF) and 23andme respiratory support summary statistics using the same methodology. Each meta-analysis summary statistics were adjusted with genomic control using METAL.

### Identification of significant and novel loci

We identified significant loci and their lead variants with a recursive procedure: (1) ranked variants by increasing *P*-value in the multi-study meta-analysis, (2) selected top variant as lead, (3) removed all variants within ±1Mbp of the top variant signal, (4) re-ranked remaining variants, (5) repeated steps 1-4 until no variants remained with *P <* 5 × 10^*−*8^. For the identified lead variants we additionally required nominal significance (*P <* 0.05) and consistency in the direction of effect with the GenOMICC severe versus ctrl-mld GWAS meta-analysis. Sentinel variants passing all these criteria are shown in Supplementary Material. We identified novel signals by filtering for genome-wide significant signals that were outside ±1Mbp of the lead variants identified by the latest critical COVID-19 meta-analyses (Pairo-Castineira et al. 2023^3^ and HGI version 7 - phenotype A2^13^. The extended MHC/HLA region (GRCh38, chr6:25,726,063-33,400,644) ^36^ was treated as a single locus.

### Fine-mapping

We used MESuSiE ^16^ to fine-map locus signals from the multi-study meta-analysis that contained genome-wide significant (*P <* 5 × 10^*−*8^) variants in the GenOMICC severe vs ctrl-all GWAS meta-analysis. This allowed us to use the LD from the same population samples as those that generated the GWAS summary statistics, increasing precision. Fine-mapping followed these steps: (1): We performed LD-clumping on GenOMICC severe vs ctrl-all ancestry-specific (AFR, EAS, EUR, SAS) and meta-analysis GWAS summary statistics; (2) we took the lead variants from LD-clumps falling within multi-study meta-analysis loci and defined regions +-500Kbp on each side of the lead variants while merging regions that overlapped; (3) we extracted pairwise LD (r) among all variants and summary statistics across AFR, EAS, EUR, SAS for each region; (4) we performed checks for LD mismatches with MESuSiE *kriging rss* function and removed SNPs that satisfied either of the following criteria: (a) *logLR ≥* 2 and |*Z*| *≥* 2, (b) *z*_*stddiff*_ *>* 3; (5) we then ran fine-mapping with MESuSiE for each region with defaults: *L*=10, min abs(*r*)=0.5, posterior probability for CS *≥* 0.95. We did not perform fine-mapping of the signal in the MHC region.

### Polygenic risk score (PRS) analysis

We calculated a Polygenic risk score (PRS) using the GenOMICC severe vs ctrl-mld cohort. We randomly allocated 80% of European individuals into a training cohort to construct the PRS and used the remaining 20% as a test cohort. We performed a GWAS with training data with the same covariates as the main GWAS. We trained the PRS with LDpred2^37^ using the bigsnpR package ^38^ and the severe vs ctrl-mld cohort European genotypes as reference panel. We calculated a null model with a logistic regression of the gwas covariates (age, sex, age square, aged by sex and 20 PCs), and we compared it to a model adding the PRS. ROC curves and AUC were calculated using the pROC R package ^39^.

### Gene-based rare variant analysis (RVAT)

#### Variant annotation and masks

We annotated all variants with variant consequences in the aggregated genomic data mild COVID-19 using VEP version 105^17^. We also annotated all variants with the Combined Annotation-Dependent Depletion pathogenicity score tool (CADD) version 1.6^40^. We filtered variant consequences for the canonical transcript. We defined two masks *strict_LoF* and *mild_LoF*. The *strict_LoF* mask included High confidence Loss of function (HC-LOF) variants as annotated by LOFTEE ^41^. The *mild_LoF* mask included the *strict_LoF* mask variants and missense and protein-altering mutations that had CADD*>*= 10.

#### Site-QC

We filtered for sites with minor allele frequency (maf) *<* 0.005 and missingness *<* 0.05. We additionally tested for differential missingness with plink v1.9 and removed variants with *P <* 10^*−*5^.

#### GenOMICC analysis

We performed rare variant analysis on a per ancestry basis using REGENIE version 3.2.5^18^. We used sex, age, age^2^, age × sex, the first 10 principal components from high quality common variants and the first 10 principal components from high quality rare variants. We performed burden, SKAT and ACAT-V tests as implemented in REGENIE and combined *P*-values across tests and allele frequency threshold classes (singletons and *<* 0.005) with a single omnibus test (ACAT-O) to maximise power.

#### Multi-study meta-analysis

We performed meta-analysis aggregating data from our results (GenOMICC severe vs ctrl-dgn) and other participating studies that matched the analysis plan and contributed summary statistics. Participating studies performed independent RVAT analysis for the GenOMICC phenotype definition (phenotype A2 in Covid-19 HGI was designed to match this), with cases being laboratory confirmed Covid-19 with one or more of the following outcomes: Death ECMO requirement Mechanical ventilation (i.e. intubation) requirement Non-invasive ventilation requirement (i.e. new requirement for BiPAP or CPAP) High-flow oxygen therapy requirement (e.g. Optiflow) and controls being every other participant in each cohort that is not a case. For meta-analysing *P*-values across studies, we used Stouffer’s weighted Z-score method and weighted with the effective sample size of each study calculated as 4*/*(1*/*(*N*_*case*_ + *N*_*control*_)).

### Structural analysis for SLC50A1

SLC50A1 (UniProt ID Q9BRV3) was modelled using AlphaFold2^42^ as implemented on the Google Colab webserver ^43^. *β*-D-glucopyranose was computationally docked to AlphaFold models of SLC50A1 using GN-INA (estimated free energy of binding *−*6.64 and *−*6.77 kcal/mol for wild type and Arg201Leu SLC50A1, respectively) as implemented on the Tamarind Bio webserver ^44^. The effect of the Arg201Leu mutation on SLC50A1 stability was calculated using the AlphaFold model of SLC50A1 and mCSM (predicted stability change ΔΔ*G −*1.379 kcal/mol, destabilizing) ^45^. Structure figures were prepared using the PyMOL Molecular Graphics System 2.6 from Schrödinger, LLC. C-terminal residues Glu211-Thr221 are not shown for simplicity in these figures.

## Data Availability

All other data produced in the present study are available upon reasonable request to the authors

https://genomicc.org/data

## Code availability

Analysis plan for rare variant analysis and code shared across participating studies is publicly available at GitHub: https://github.com/genomicsengland/COVID19_GenOMICC_AVT_analysis.

## Acknowledgements

We thank the patients and their loved ones who volunteered to contribute to this study, and the research staff in every intensive care unit who recruited patients. GenOMICC was funded by Sepsis Research (the Fiona Elizabeth Agnew Trust), the Intensive Care Society, a Wellcome Trust Senior Research Fellowship (J.K.Baillie, 223164/Z/21/Z), the Department of Health and Social Care (DHSC), Illumina, LifeArc, the Medical Research Council, UKRI, a BBSRC Institute Strategic Program Support Grant to the Roslin Institute (BBS/E/D/20002172, BBS/E/D/10002070 and BBS/E/D/30002275) and UKRI grants MC PC 20004, MC PC 19025, MC PC 1905, and MRNO2995X/1. ADB acknowledges funding from the Wellcome PhD training fellowship for clinicians (204979/Z/16/Z), the Edinburgh Clinical Academic Track (ECAT) programme. This research is supported in part by the Data and Connectivity National Core Study, led by Health Data Research UK in partnership with the Office for National Statistics and funded by UK Research and Innovation (grant ref MC PC 20029). This study owes a great deal to the National Institute for Healthcare Research Clinical Research Network (NIHR CRN) and the Chief Scientist’s Office (Scotland), who facilitate recruitment into research studies in NHS hospitals, and to the global ISARIC and InFACT consortia. This work forms part of the translational research portfolio of the National Institute for Health and Care Research Barts Biomedical Research Centre. T.M. is supported by Cancer Research UK grant DRCRPG-May23/100002 to C. Siebold. Genomics England: This research was made possible through access to data in the National Genomic Research Library ^46^, which is managed by Genomics England Limited (a wholly owned company of the Department of Health and Social Care). The National Genomic Research Library (https://www.genomicsengland.co.uk/research) holds data provided by patients and collected by the NHS as part of their care and data collected as part of their participation in research. The National Genomic Research Library is funded by the National Institute for Health Research and NHS England. The Wellcome Trust, Cancer Research UK and the Medical Research Council have also funded research infrastructure. REACT: National Institute for Health and Care Research (NIHR) and UK Research and Innovation (UKRI) - REACT-Genomics England (REACT-GE) (MR/V030841/1) and REACT-Long COVID (REACT-LC) (COV-LT-0040). The REACT study was funded by the UK Department of Health and Social Care with supplemental funding from the Huo Family Foundation. For the purpose of open access, the author has applied a CC BY public copyright licence to any Author Accepted Manuscript version arising from this submission.

