## Supplementary Information for "Whole genome sequence meta-analyses reveal common and rare genetic associations with critical COVID-19"

### Supplementary Material

**2 Additional figures** **23**

**3 Acknowledgments** **32**

**4 Conflicts of Interest** **33**

**5 Extended References** **33**

### 1 Supplementary note

This section presents additional analyses and detailed results that complement the main findings.

#### 1.1 Genome-wide association study analyses (GWAS)

##### 1.1.1 GenOMICC cohorts, multi-ancestry GWAS and meta-analysis

Following stringent quality control procedures (Methods), we conducted Genome-wide association analyses (GWAS) using SAIGE[?] for a cohort (GenOMICC) of 11,423 critically ill cases and 60,628 controls. 9,357 (82%) of the cases were part of the primary analysis in our prior publication[1]. The controls included 100,000 Genomes Project (100kGP) individuals (n = 49,360) and recruited individuals with mild COVID-19 (n = 11,268) (Methods), mainly as part of the Real-time assessment of community transmission (REACT) study (cohort breakdown by ancestry shown in Extended Data Fig. 1).

GWAS analyses were carried out separately for four genetically inferred ancestry groups (African (AFR); East Asian (EAS); European (EUR); South Asian (SAS)) and using either all controls (ctrl-all) or only individuals with mild COVID-19 (ctrl-mld) with cohort breakdowns by ancestry shown in Extended Data Fig. 1. Demographic characteristics in terms of age and sex were broadly matched between mild COVID-19 and 100kGP control cohorts (Figure 1, Table 1). Severe cases and controls (ctrl-all) were broadly matched in terms of their ancestry composition (Figure 2). GWAS results were meta-analysed across ancestries and for each set of controls separately using inverse-variance-weighted fixed effects meta-analysis (IVW) (Methods).

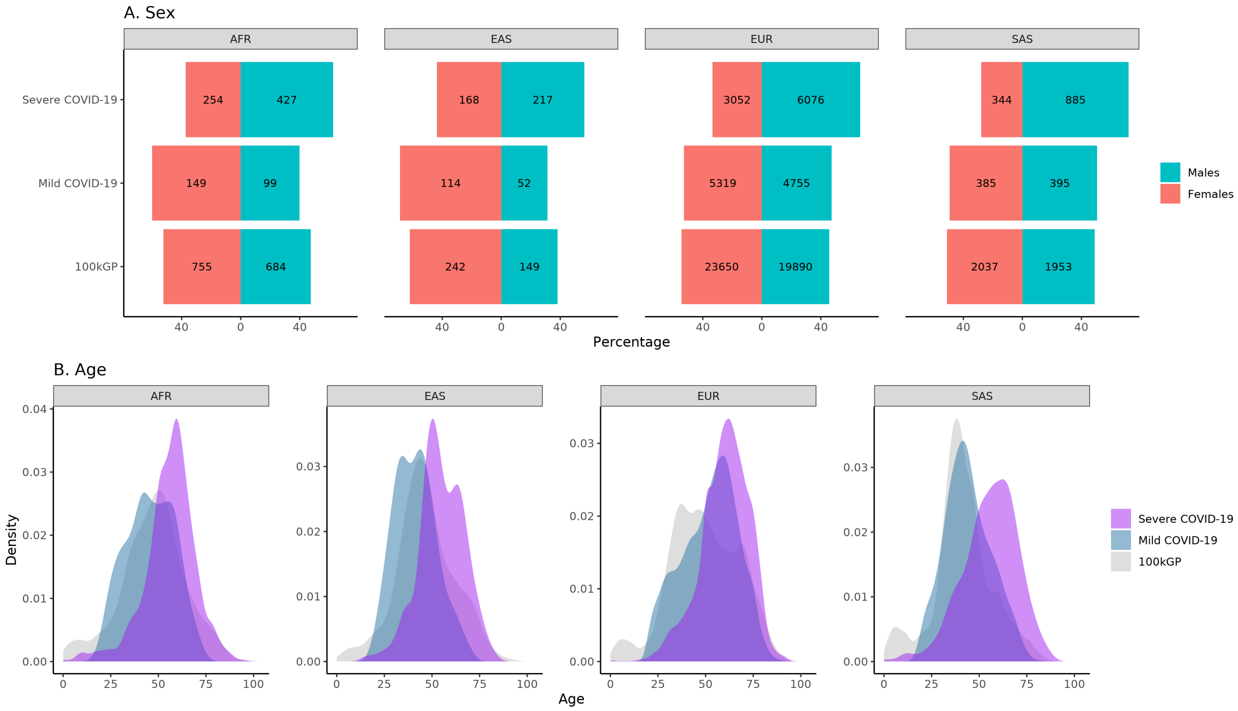

Figure 1: Demographic characteristics of analysed cohorts for the GenOMICC study

| Cohort | All ancestries | AFR | EAS | EUR | SAS |
|---|---|---|---|---|---|
| All | 51[26] | 51[20] | 49[19] | 52[26] | 43[20] |
| Severe COVID-19 | 60[17] | 57.6[14] | 54[15.8] | 61[17] | 57[19] |
| Mild COVID-19 | 53[22] | 46[20] | 41[16] | 54[21.8] | 43[17.2] |
| 100kGP | 48[26] | 48[21] | 45[18] | 49[26] | 41[15] |

Table 1: Age medians and interquartile range in brackets across cohorts and ancestries.

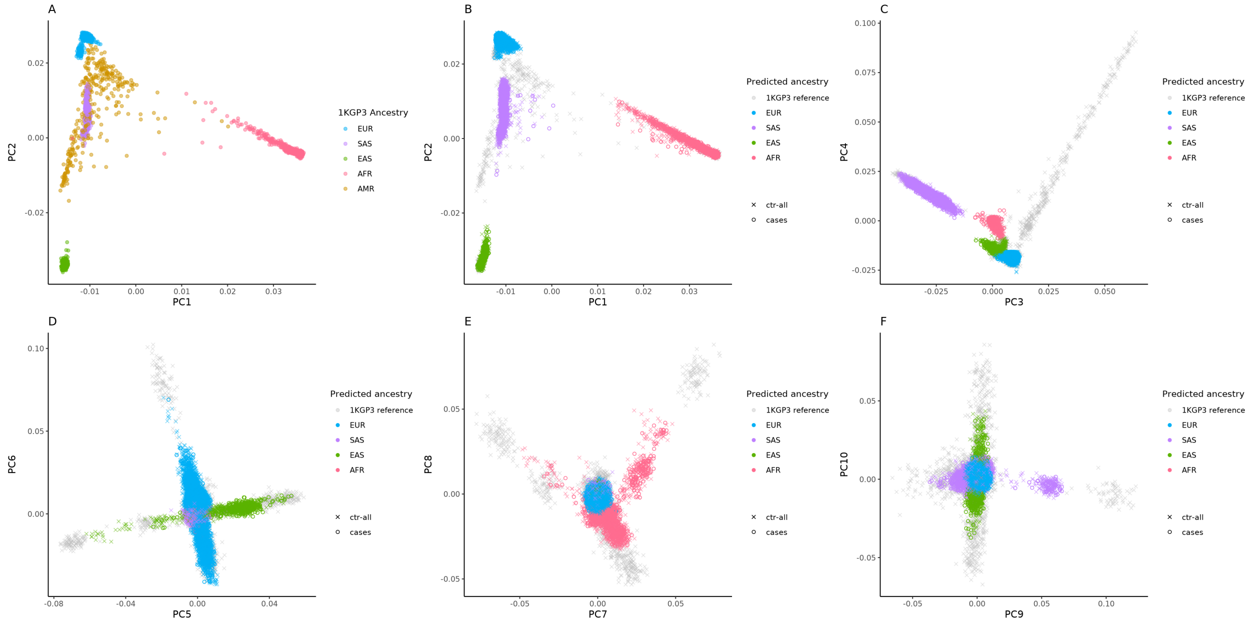

Figure 2: Projection of severe cases vs. ctrl-all samples onto 1,000 Genomes Project (1KGP3) principal components with coloring by predicted ancestry. Panel A shows the first two PCs for the 1KGP3 samples. Panels B-F show projections of the severe cases and ctrl-all samples of this study coloured by predicted ancestry, with PCs for 1KGP3 samples shown in grey.

##### 1.1.2 Multi-study GWAS meta-analysis

To maximise power for GWAS discovery, we meta-analysed the GWAS summary statistics from three studies: GenOMICC severe versus ctrl-all (this study), HGIv7-A2_ALL_leave_23andme_and_genomicc[2], and 23andMe EUR respiratory support[3] (included cohorts breakdown in Extended Data Fig. 1). To account for potential heterogeneity in variant overlap and ancestry composition across studies that could confound conditional analysis on the basis of meta-analysis summaries[4], we defined locus associations as ±1Mbp regions surrounding top variant signals (Methods). We identified 38 genome-wide significant loci ($P < 5 \times 10^{-8}$), of which five were not previously reported, near *ARF1*, *MECOM*, *ZNF462*, *KLF13* and *MVP* genes (summary statistics in Table 2, allele frequencies in Table **??**)

Each of the novel lead variant signals for *ARF1*, *MECOM*, *ZNF462*, *KLF13* was supported by multiple genome-wide significant variants ($P < 5 \times 10^{-8}$) in strong LD with the lead (EUR $r^2_{EUR} > 0.8$; Figure 3). *MVP* was supported by three variants with $P < 10^{-4}$ in moderate LD with the lead ($r^2_{AFR} > 0.4$; Figure 3). Additionally, lead variant odds ratios for all signals were consistent with meta-analysis using only the ctrl-mld subset of controls (Table 4, Figure 4, and Figure 3).

We further assessed between-study heterogeneity and support from the second largest study (HGI) to increase confidence in these associations. For the latter, we considered a $P < 0.0125$ (taken as $P = 0.05$ corrected for the number of assessed signals) with the same direction of effect in the HGI cohort as evidence supporting the respective signal. Signals near *ARF1*, *ZNF462*, *KLF13* had support across studies and no evidence of heterogeneity, but *MECOM* has $P > 0.0125$ in the HGI cohort and $P_{het} = 2.13 \times 10^{-3}$ (Table 1, main manuscript). We therefore consider the signal near *MECOM* as requiring further replication in an independent cohort.

The locus near *MVP* could not be assessed because no other studies had sufficient data. The lead variant is very rare in non-African ancestries (maf of $1.3 \times 10^{-4}$ in gnomAD NFE, v4.1).

| Chr:pos(b38):ref:alt | rsid | $OR$ | $OR_{CI}$ | $P_{meta}$ | $P_{het}$ | $P_{GenOMICC}$ | $P_{hgi}$ | Nearest gene | Citation |
|---|---|---|---|---|---|---|---|---|---|
| 1:9062610:A:G | rs2505973 | 0.932 | 0.91-0.954 | $4.00 \times 10^{-9}$ | $4.32 \times 10^{-1}$ | $3.82 \times 10^{-7}$ | $1.63 \times 10^{-3}$ | *SLC2A5* | GenOMICC3 |
| 1:64951326:A:G* | rs12730021 | 0.897 | 0.871-0.923 | $3.90 \times 10^{-13}$ | $3.52 \times 10^{-1}$ | $2.54 \times 10^{-10}$ | $1.54 \times 10^{-4}$ | *JAK1* | GenOMICC3 |
| 1:155197995:A:G* | rs41264915 | 0.823 | 0.793-0.854 | $8.62 \times 10^{-25}$ | $5.08 \times 10^{-1}$ | $1.91 \times 10^{-16}$ | $9.71 \times 10^{-10}$ | *THBS3* | GenOMICC2 |
| 1:228089434:C:T | rs3738681 | 0.881 | 0.843-0.92 | $9.30 \times 10^{-9}$ | $2.55 \times 10^{-1}$ | $1.00 \times 10^{-5}$ | $1.10 \times 10^{-3}$ | *ARF1* | Novel |
| 2:60480453:A:G* | rs1123573 | 0.904 | 0.882-0.926 | $9.77 \times 10^{-16}$ | $3.22 \times 10^{-1}$ | $1.41 \times 10^{-9}$ | $2.81 \times 10^{-8}$ | *BCL11A* | GenOMICC2 |
| 3:45818159:G:A* | rs17713054 | 2.05 | 1.97-2.14 | $8.34 \times 10^{-266}$ | $3.49 \times 10^{-10}$ | $2.18 \times 10^{-192}$ | $1.14 \times 10^{-77}$ | *LZTFL1* | SCGG19 |
| 3:146517122:G:A* | rs343320 | 1.15 | 1.1-1.21 | $1.30 \times 10^{-9}$ | $7.23 \times 10^{-2}$ | $1.72 \times 10^{-9}$ | $3.38 \times 10^{-2}$ | *PLSCR1* | GenOMICC2 |
| 3:169081115:G:GGAT* | rs112039646 | 1.15 | 1.1-1.2 | $4.81 \times 10^{-9}$ | - | $4.58 \times 10^{-9}$ | - | *MECOM* | Novel |
| 4:25446871:A:G | rs7664615 | 0.906 | 0.879-0.933 | $3.27 \times 10^{-11}$ | $2.59 \times 10^{-1}$ | $2.39 \times 10^{-7}$ | $3.11 \times 10^{-4}$ | *ANAPC4* | GenOMICC3 |
| 4:105897896:G:A* | rs34712979 | 0.902 | 0.875-0.929 | $9.31 \times 10^{-12}$ | $2.12 \times 10^{-1}$ | $2.23 \times 10^{-9}$ | $4.15 \times 10^{-4}$ | *NPNT* | GenOMICC3 |
| 5:132422622:A:G* | rs2269821 | 0.908 | 0.879-0.938 | $5.21 \times 10^{-9}$ | $2.27 \times 10^{-1}$ | $1.56 \times 10^{-7}$ | $1.86 \times 10^{-3}$ | *IRF1* | GenOMICC2 |
| 6:31182658:A:G | rs1128175 | 0.889 | 0.865-0.914 | $1.11 \times 10^{-16}$ | $2.15 \times 10^{-1}$ | $3.80 \times 10^{-8}$ | $1.98 \times 10^{-8}$ | *HLA* | GenOMICC1 |
| 6:41520640:G:A* | rs12660421 | 1.41 | 1.33-1.5 | $7.99 \times 10^{-29}$ | $2.24 \times 10^{-1}$ | $2.26 \times 10^{-18}$ | $2.38 \times 10^{-11}$ | *FOXP4* | HGI21 |
| 7:75634474:C:T | rs12534422 | 1.08 | 1.05-1.11 | $9.90 \times 10^{-10}$ | $6.37 \times 10^{-1}$ | $5.56 \times 10^{-7}$ | $5.78 \times 10^{-4}$ | *HIP1* | GenOMICC3 |
| 7:100032719:C:T* | rs2897075 | 1.09 | 1.06-1.11 | $7.31 \times 10^{-13}$ | $6.55 \times 10^{-1}$ | $2.15 \times 10^{-8}$ | $1.35 \times 10^{-5}$ | *ZKSCAN1* | GenOMICC3 |
| 8:60568368:GTC:G | rs34038069 | 1.07 | 1.05-1.1 | $7.00 \times 10^{-9}$ | $5.10 \times 10^{-1}$ | $1.84 \times 10^{-6}$ | $7.13 \times 10^{-4}$ | *RAB2A* | GenOMICC3 |
| 9:21206606:C:G* | rs28368148 | 1.54 | 1.38-1.73 | $7.95 \times 10^{-14}$ | $2.76 \times 10^{-1}$ | $4.51 \times 10^{-11}$ | $1.10 \times 10^{-4}$ | *IFNA10* | GenOMICC2 |
| 9:106709885:G:A | rs60568503 | 1.07 | 1.05-1.1 | $1.28 \times 10^{-8}$ | $6.97 \times 10^{-1}$ | $4.65 \times 10^{-6}$ | $4.61 \times 10^{-4}$ | *ZNF462* | Novel |
| 9:133271182:T:C* | rs550057 | 0.889 | 0.866-0.912 | $1.04 \times 10^{-18}$ | $7.73 \times 10^{-1}$ | $7.57 \times 10^{-9}$ | $1.24 \times 10^{-10}$ | *ABO* | SCGG19 |
| 10:79946568:A:G* | rs721917 | 1.09 | 1.06-1.11 | $1.87 \times 10^{-13}$ | $1.01 \times 10^{-1}$ | $4.00 \times 10^{-8}$ | $1.77 \times 10^{-5}$ | *SFTPD* | HGI22 |
| 11:1214934:A:G | rs12802931 | 0.899 | 0.87-0.929 | $1.54 \times 10^{-10}$ | $7.17 \times 10^{-1}$ | $1.98 \times 10^{-6}$ | $1.15 \times 10^{-5}$ | *MUC5B* | HGI22 |
| 11:34482745:G:A* | rs61882275 | 0.885 | 0.864-0.906 | $2.29 \times 10^{-24}$ | $2.44 \times 10^{-1}$ | $3.42 \times 10^{-17}$ | $2.85 \times 10^{-9}$ | *ELF5* | GenOMICC2 |
| 12:112919388:G:A* | rs10774671 | 1.11 | 1.08-1.14 | $9.89 \times 10^{-16}$ | $9.61 \times 10^{-1}$ | $1.06 \times 10^{-9}$ | $1.35 \times 10^{-7}$ | *OAS1* | GenOMICC1 |
| 12:132481571:G:A* | rs11614702 | 1.11 | 1.08-1.13 | $3.71 \times 10^{-18}$ | $6.04 \times 10^{-2}$ | $1.27 \times 10^{-14}$ | $2.36 \times 10^{-6}$ | *FBRSL1* | GenOMICC2 |
| 13:112881427:C:T* | rs12585036 | 1.14 | 1.11-1.17 | $1.14 \times 10^{-20}$ | $9.50 \times 10^{-2}$ | $3.87 \times 10^{-16}$ | $6.72 \times 10^{-7}$ | *ATP11A* | GenOMICC2 |
| 15:31319838:G:T* | rs11636034 | 0.924 | 0.9-0.948 | $1.30 \times 10^{-9}$ | $1.24 \times 10^{-1}$ | $3.68 \times 10^{-8}$ | $2.47 \times 10^{-3}$ | *KLF13* | Novel |
| 16:29843264:G:A* | rs138640006 | 2.22 | 1.67-2.94 | $2.66 \times 10^{-8}$ | - | $2.53 \times 10^{-8}$ | - | *MVP* | Novel |
| 16:89196249:G:A* | rs117169628 | 1.16 | 1.12-1.2 | $3.07 \times 10^{-17}$ | $2.70 \times 10^{-1}$ | $2.44 \times 10^{-9}$ | $1.26 \times 10^{-7}$ | *SLC22A31* | GenOMICC2 |
| 17:40003082:T:C* | rs12941811 | 1.08 | 1.05-1.1 | $3.82 \times 10^{-10}$ | $3.50 \times 10^{-2}$ | $1.89 \times 10^{-8}$ | $7.05 \times 10^{-3}$ | *PSMD3* | GenOMICC3 |
| 17:46779515:G:A | rs199514 | 1.11 | 1.08-1.15 | $9.70 \times 10^{-13}$ | $5.95 \times 10^{-1}$ | $5.35 \times 10^{-6}$ | $2.31 \times 10^{-8}$ | *WNT3* | Degenhardt |
| 17:49863260:C:A* | rs3848456 | 1.32 | 1.24-1.4 | $8.79 \times 10^{-20}$ | $3.92 \times 10^{-1}$ | $8.95 \times 10^{-12}$ | $4.37 \times 10^{-10}$ | *TAC4* | Degenhardt |
| 19:4717660:A:G* | rs12610495 | 1.26 | 1.23-1.29 | $6.13 \times 10^{-67}$ | $2.59 \times 10^{-2}$ | $1.70 \times 10^{-48}$ | $6.52 \times 10^{-22}$ | *DPP9* | GenOMICC1 |
| 19:10352442:G:C* | rs34536443 | 1.48 | 1.39-1.58 | $4.69 \times 10^{-35}$ | $5.93 \times 10^{-1}$ | $6.23 \times 10^{-25}$ | $2.05 \times 10^{-10}$ | *TYK2* | GenOMICC1 |
| 19:48702888:G:C* | rs516316 | 0.914 | 0.893-0.936 | $2.50 \times 10^{-14}$ | $8.50 \times 10^{-2}$ | $3.69 \times 10^{-12}$ | $1.28 \times 10^{-4}$ | *FUT2* | GenOMICC2 |
| 20:6489447:G:A | rs2326788 | 0.928 | 0.905-0.951 | $2.14 \times 10^{-9}$ | $9.06 \times 10^{-1}$ | $7.04 \times 10^{-6}$ | $1.89 \times 10^{-4}$ | *BMP2* | GenOMICC3 |
| 21:33240996:A:G* | rs17860169 | 1.2 | 1.17-1.23 | $1.83 \times 10^{-49}$ | $1.80 \times 10^{-1}$ | $2.15 \times 10^{-32}$ | $6.18 \times 10^{-19}$ | *IFNAR2* | GenOMICC1 |
| 21:41471061:G:A* | rs9305744 | 0.909 | 0.885-0.934 | $5.48 \times 10^{-12}$ | $7.83 \times 10^{-1}$ | $4.13 \times 10^{-8}$ | $3.85 \times 10^{-5}$ | *TMPRSS2* | GenOMICC3 |
| X:15602217:T:C* | rs190509934 | 0.465 | 0.373-0.58 | $1.02 \times 10^{-11}$ | $4.45 \times 10^{-1}$ | $2.92 \times 10^{-9}$ | $5.26 \times 10^{-4}$ | *ACE2* | HGI22 |

Table 2: Lead variants of locus association signals in the multi-study GWAS meta-analysis. Variant ids are provided as Chr:pos(b38):ref:alt using GRCh38 coordinates with ref indicating the reference allele and alt being the effect allele and an asterisk (*) indicates that the locus indexed by the lead variant is fine-mapped. $OR$, $OR_{CI}$ and $P_{meta}$ are Odds ratios, Odds ratio 95% confidence intervals, and the $P$-value from the meta-analysis. $P_{het}$ is the $P$-value from heterogeneity analysis. $P_{GenOMICC}$ and $P_{hgi}$ are $P$-values for lead variants from the GenOMICC multi-ancestry meta-analysis and HGIv7-leaveGenOMICC data, respectively. Citation column refers to the first study reporting a significant signal within ±1Mbp of the lead variant of this study, with "Novel" indicating novel significant loci identified in this study (Referenced studies being either GenOMICC1[1], GenOMICC2[5], GenOMICC3[1], SCGG19[6], HGI21[7] , HGI22[8], Degenhardt[9]). Signals within the extended MHC region are annotated as "HLA".

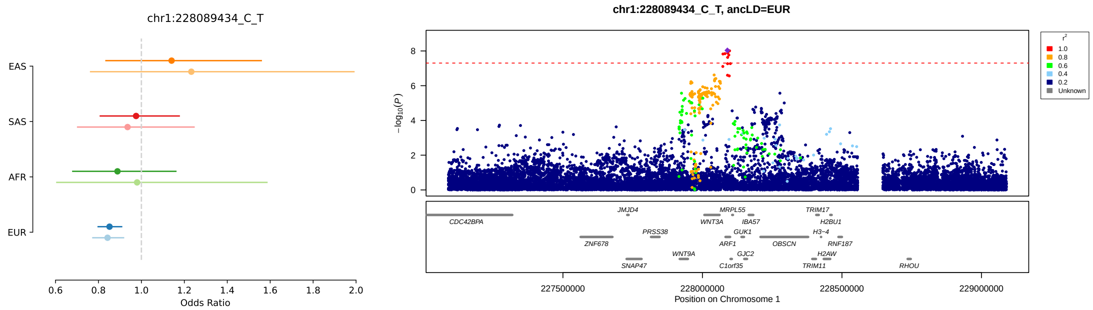
chr1:228089434_C_T
EAS
SAS
AFR
EUR
0.6 0.8 1.0 1.2 1.4 1.6 1.8 2.0
Odds Ratio
chr1:228089434_C_T, ancLD=EUR
−log10(P)
r²
1.0
0.8
0.6
0.4
0.2
Unknown
CDC42BPA
JMJD4
ZNF678
PRSS38
SNAP47
WNT9A
MRPL55
WNT3A
ARF1
GUK1
C1orf35
IBA57
GJC2
OBSCN
TRIM17
H2BU1
H3−4
RNF187
H2AW
TRIM11
RHOU
227500000 228000000 228500000 229000000
Position on Chromosome 1

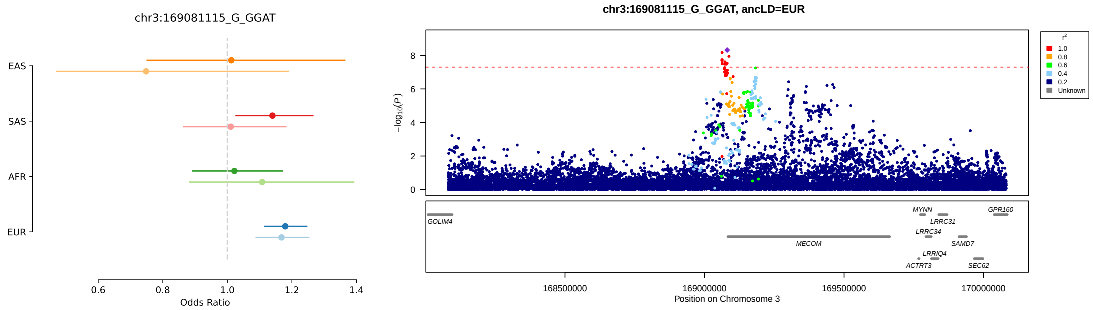
chr3:169081115_G_GGAT
EAS
SAS
AFR
EUR
0.6 0.8 1.0 1.2 1.4
Odds Ratio
chr3:169081115_G_GGAT, ancLD=EUR
−log10(P)
r²
1.0
0.8
0.6
0.4
0.2
Unknown
GOLIM4
MECOM
MYNN
LRRC31
LRRC34
LRRIQ4
ACTRT3
SAMD7
GPR160
SEC62
168500000 169000000 169500000 170000000
Position on Chromosome 3

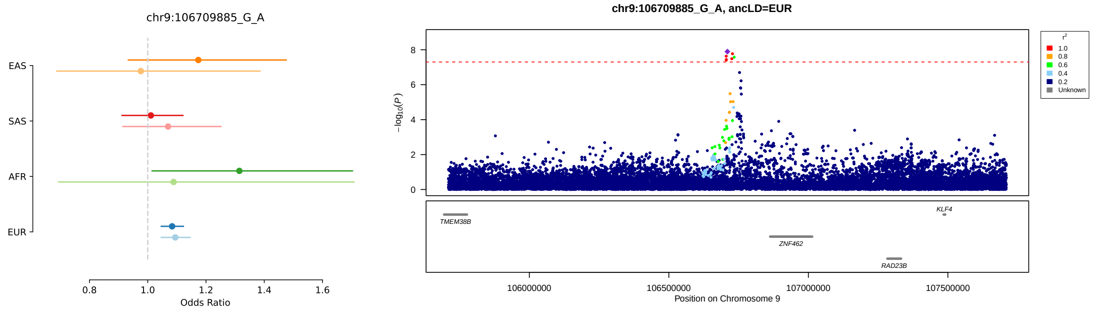
chr9:106709885_G_A
EAS
SAS
AFR
EUR
0.8 1.0 1.2 1.4 1.6
Odds Ratio
chr9:106709885_G_A, ancLD=EUR
−log10(P)
r²
1.0
0.8
0.6
0.4
0.2
Unknown
TMEM38B
ZNF462
RAD23B
KLF4
106000000 106500000 107000000 107500000
Position on Chromosome 9

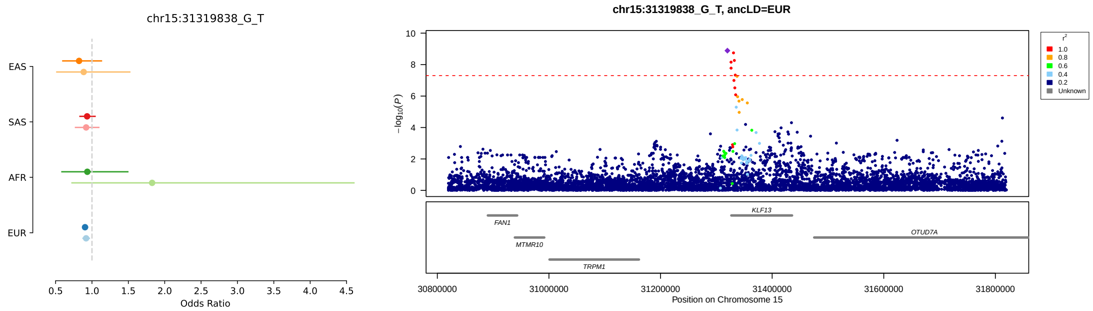
chr15:31319838_G_T
EAS
SAS
AFR
EUR
0.5 1.0 1.5 2.0 2.5 3.0 3.5 4.0 4.5
Odds Ratio
chr15:31319838_G_T, ancLD=EUR
−log10(P)
r²
1.0
0.8
0.6
0.4
0.2
Unknown
FAN1
MTMR10
TRPM1
KLF13
OTUD7A
30800000 31000000 31200000 31400000 31600000 31800000
Position on Chromosome 15

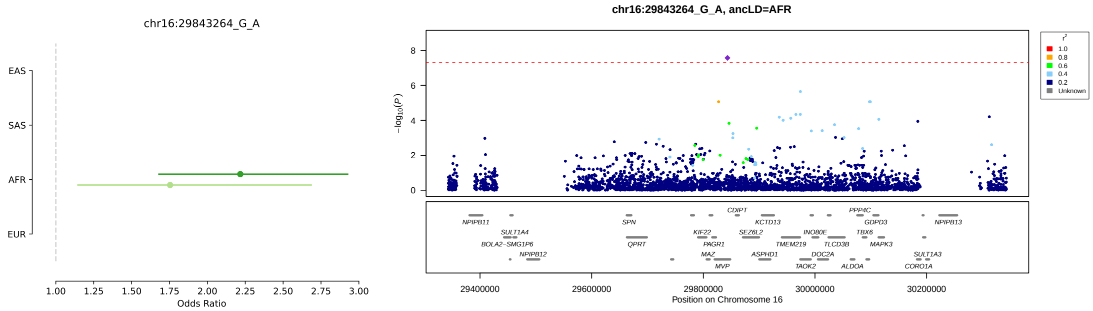

Figure 3: LocusZoom and forest plots illustrating the novel genetic association signals identified in this study. Panels on the right compare cross-ancestry odds ratios of lead variants from analyses using all controls (ctrl-all, darker lines) vs. analyses using only controls with mild COVID-19 (ctrl-mld, lighter lines). Panels on the right show LocusZoom plots centered on lead variant. $P$-values are from the cross-ancestry GenOMICC severe versus ctrl-all meta-analysis. The population used to calculate LD is indicated on the title and corresponds to the population having the smallest $P$-value in the GenOMICC severe vs ctrl-all meta-analysis. Equivalent plots for all lead variants in Table 2 are in Figure 9.

| Chr:pos(b38):ref:alt | rsid | $OR$ | $OR_{CI}$ | $P$ | $OR_{ctr-mld}$ | $OR_{CI_{ctr-mld}}$ | $P_{ctr-mld}$ |
|---|---|---|---|---|---|---|---|
| 1:9062610:A:G | rs2505973 | 0.932 | 0.91-0.954 | $4.00 \times 10^{-9}$ | 0.906 | 0.867-0.946 | $8.99 \times 10^{-6}$ |
| 1:64951326:A:G | rs12730021 | 0.897 | 0.871-0.923 | $3.90 \times 10^{-13}$ | 0.931 | 0.884-0.981 | $7.49 \times 10^{-3}$ |
| 1:155197995:A:G | rs41264915 | 0.823 | 0.793-0.854 | $8.62 \times 10^{-25}$ | 0.757 | 0.707-0.811 | $2.66 \times 10^{-15}$ |
| 1:228089434:C:T | rs3738681 | 0.881 | 0.843-0.92 | $9.30 \times 10^{-9}$ | 0.861 | 0.794-0.934 | $2.84 \times 10^{-4}$ |
| 2:60480453:A:G | rs1123573 | 0.904 | 0.882-0.926 | $9.77 \times 10^{-16}$ | 0.919 | 0.879-0.96 | $1.66 \times 10^{-4}$ |
| 3:45818159:G:A | rs17713054 | 2.05 | 1.97-2.14 | $8.34 \times 10^{-266}$ | 2.06 | 1.93-2.2 | $2.36 \times 10^{-100}$ |
| 3:146517122:G:A | rs343320 | 1.15 | 1.1-1.21 | $1.30 \times 10^{-9}$ | 1.17 | 1.08-1.26 | $7.58 \times 10^{-5}$ |
| 3:169081115:G:GGAT | rs112039646 | 1.15 | 1.1-1.2 | $4.81 \times 10^{-9}$ | 1.13 | 1.06-1.2 | $1.17 \times 10^{-4}$ |
| 4:25446871:A:G | rs7664615 | 0.906 | 0.879-0.933 | $3.27 \times 10^{-11}$ | 0.897 | 0.851-0.945 | $4.44 \times 10^{-5}$ |
| 4:105897896:G:A | rs34712979 | 0.902 | 0.875-0.929 | $9.31 \times 10^{-12}$ | 0.916 | 0.87-0.964 | $7.60 \times 10^{-4}$ |
| 5:132422622:A:G | rs2269821 | 0.908 | 0.879-0.938 | $5.21 \times 10^{-9}$ | 0.882 | 0.829-0.94 | $9.27 \times 10^{-5}$ |
| 6:31182658:A:G | rs1128175 | 0.889 | 0.865-0.914 | $1.11 \times 10^{-16}$ | 0.916 | 0.871-0.963 | $5.40 \times 10^{-4}$ |
| 6:41520640:G:A | rs12660421 | 1.41 | 1.33-1.5 | $7.99 \times 10^{-29}$ | 1.43 | 1.27-1.61 | $4.88 \times 10^{-9}$ |
| 7:75634474:C:T | rs12534422 | 1.08 | 1.05-1.11 | $9.90 \times 10^{-10}$ | 1.1 | 1.05-1.15 | $5.44 \times 10^{-5}$ |
| 7:100032719:C:T | rs2897075 | 1.09 | 1.06-1.11 | $7.31 \times 10^{-13}$ | 1.08 | 1.03-1.13 | $7.37 \times 10^{-4}$ |
| 8:60568368:GTC:G | rs34038069 | 1.07 | 1.05-1.1 | $7.00 \times 10^{-9}$ | 1.04 | 1-1.09 | $4.40 \times 10^{-2}$ |
| 9:21206606:C:G | rs28368148 | 1.54 | 1.38-1.73 | $7.95 \times 10^{-14}$ | 1.58 | 1.32-1.89 | $7.35 \times 10^{-7}$ |
| 9:106709885:G:A | rs60568503 | 1.07 | 1.05-1.1 | $1.28 \times 10^{-8}$ | 1.09 | 1.04-1.14 | $9.34 \times 10^{-5}$ |
| 9:133271182:T:C | rs550057 | 0.889 | 0.866-0.912 | $1.04 \times 10^{-18}$ | 0.936 | 0.892-0.982 | $7.47 \times 10^{-3}$ |
| 10:79946568:A:G | rs721917 | 1.09 | 1.06-1.11 | $1.87 \times 10^{-13}$ | 1.08 | 1.04-1.13 | $2.03 \times 10^{-4}$ |
| 11:1214934:A:G | rs12802931 | 0.899 | 0.87-0.929 | $1.54 \times 10^{-10}$ | 0.904 | 0.855-0.956 | $3.98 \times 10^{-4}$ |
| 11:34482745:G:A | rs61882275 | 0.885 | 0.864-0.906 | $2.29 \times 10^{-24}$ | 0.88 | 0.842-0.919 | $9.98 \times 10^{-9}$ |
| 12:112919388:G:A | rs10774671 | 1.11 | 1.08-1.14 | $9.89 \times 10^{-16}$ | 1.12 | 1.07-1.17 | $1.18 \times 10^{-6}$ |
| 12:132481571:G:A | rs11614702 | 1.11 | 1.08-1.13 | $3.71 \times 10^{-18}$ | 1.13 | 1.08-1.18 | $2.03 \times 10^{-8}$ |
| 13:112881427:C:T | rs12585036 | 1.14 | 1.11-1.17 | $1.14 \times 10^{-20}$ | 1.17 | 1.11-1.23 | $2.84 \times 10^{-9}$ |
| 15:31319838:G:T | rs11636034 | 0.924 | 0.9-0.948 | $1.30 \times 10^{-9}$ | 0.921 | 0.879-0.964 | $4.58 \times 10^{-4}$ |
| 16:29843264:G:A | rs138640006 | 2.22 | 1.67-2.94 | $2.66 \times 10^{-8}$ | 1.75 | 1.14-2.71 | $1.11 \times 10^{-2}$ |
| 16:89196249:G:A | rs117169628 | 1.16 | 1.12-1.2 | $3.07 \times 10^{-17}$ | 1.12 | 1.06-1.19 | $1.34 \times 10^{-4}$ |
| 17:40003082:T:C | rs12941811 | 1.08 | 1.05-1.1 | $3.82 \times 10^{-10}$ | 1.06 | 1.01-1.1 | $1.57 \times 10^{-2}$ |
| 17:46779515:G:A | rs199514 | 1.11 | 1.08-1.15 | $9.70 \times 10^{-13}$ | 1.06 | 1-1.12 | $4.53 \times 10^{-2}$ |
| 17:49863260:C:A | rs3848456 | 1.32 | 1.24-1.4 | $8.79 \times 10^{-20}$ | 1.22 | 1.1-1.36 | $2.24 \times 10^{-4}$ |
| 19:4717660:A:G | rs12610495 | 1.26 | 1.23-1.29 | $6.13 \times 10^{-67}$ | 1.27 | 1.21-1.33 | $1.01 \times 10^{-25}$ |
| 19:10352442:G:C | rs34536443 | 1.48 | 1.39-1.58 | $4.69 \times 10^{-35}$ | 1.48 | 1.34-1.62 | $4.23 \times 10^{-16}$ |
| 19:48702888:G:C | rs516316 | 0.914 | 0.893-0.936 | $2.50 \times 10^{-14}$ | 0.898 | 0.861-0.938 | $8.94 \times 10^{-7}$ |
| 20:6489447:G:A | rs2326788 | 0.928 | 0.905-0.951 | $2.14 \times 10^{-9}$ | 0.948 | 0.907-0.99 | $1.62 \times 10^{-2}$ |
| 21:33240996:A:G | rs17860169 | 1.2 | 1.17-1.23 | $1.83 \times 10^{-49}$ | 1.23 | 1.17-1.28 | $1.70 \times 10^{-19}$ |
| 21:41471061:G:A | rs9305744 | 0.909 | 0.885-0.934 | $5.48 \times 10^{-12}$ | 0.892 | 0.848-0.937 | $6.94 \times 10^{-6}$ |
| X:15602217:T:C | rs190509934 | 0.465 | 0.373-0.58 | $1.02 \times 10^{-11}$ | 0.239 | 0.105-0.543 | $6.36 \times 10^{-4}$ |

Table 4: Comparison of effect sizes of lead variants from the multi-study meta-analysis versus a multi-ancestry meta-analysis of GenOMICC cases using only individuals with mild COVID-19 as controls (GenOMICC severe versus ctrl-mld). Variant ids are provided as Chr:pos(b38):ref:alt using GRCh38 coordinates with ref indicating the reference allele and alt being the effect allele. $OR$, $OR_{CI}$ and $P$ are Odds ratios, Odds ratio 95% confidence intervals and $P$-values for the multi-study meta analysis. $OR$, $OR_{CI}$ and $P$ with subscript $ctrl - mld$ are derived from the meta-analysis of the GenOMICC severe vs. ctrl-mld GWAS.

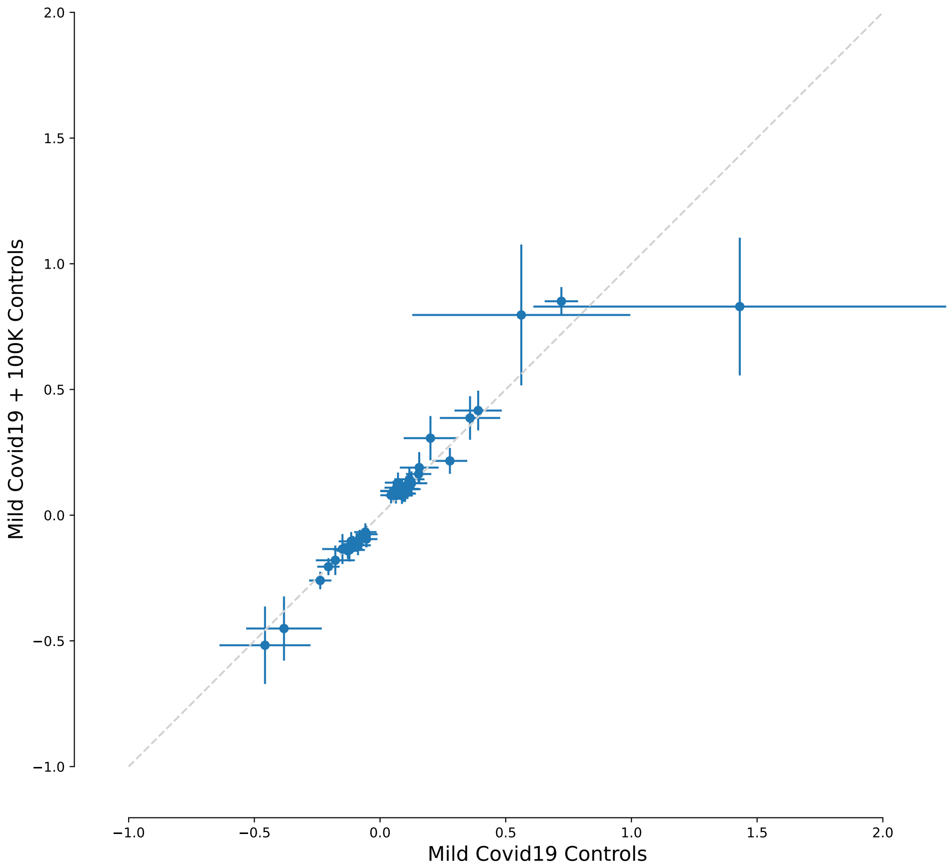

Figure 4: Comparison of logOR estimates at lead variants using either only mild Covid19 cases as controls (GenOMICC severe versus ctrl-mld) or, as in the main analysis, both mild Covid19 cases and 100K Genomes participants as controls (GenOMICC severe versus ctrl-all).

###### 1.1.3 GenOMICC multi-ancestry fine-mapping and annotation

We performed multi-ancestry fine-mapping to identify credible sets (CS) with MESuSiE[10] for a subset of 28 loci that contained significant variant signals ($P < 5 \times 10^{-8}$) in the GenOMICC severe versus ctrl-all analysis (loci indicated with asterisk on variant ID index; Table 2). We fine-mapped a total of 64 CS that were distributed within the 28 locus signals (Table 5). We annotated each CS lead variant with the best scoring V2G gene from the OpenTargets Genetics Platform and VEP predicted consequence for the canonical transcript (Table 5). Among the list of predicted consequences, we identified missense variants for *PLSCR1*, *IFNA10*, *SLC22A31* and *TYK2* genes (Table 5). We annotated additional consequences for CS variants by identifying the worst predicted VEP consequence across the CS to find an additional two missense variants at *THBS3* and *GSDMA* genes (Table 6). We also performed fine-mapping for the EUR ancestry GenOMICC cohort using SUSIE[11] which yielded a total of 58 credible sets that largely matched the multi-ancestry fine-mapping results in terms of prioritised genes and consequences (Table 7). EUR fine-mapping identified an additional missense variant for *IFNAR2* (Table 8).

| Locus index variant | iCS | nCS | CS lead variant | PIP | MinP | MinP pop | V2G gene | CADD | VEP consq | VEP gene |
|---|---|---|---|---|---|---|---|---|---|---|
| 1:64951326:A:G | 1 | 1 | 1:64951326:A:G | 1 | $8.38 \times 10^{-11}$ | META | *JAK1* | 2.3 | intron | *JAK1* |
| 1:64951326:A:G | 2 | 2 | 1:64964168:T:C | 0.57 | $2.68 \times 10^{-7}$ | META | *JAK1* | 7 | intron | *JAK1* |
| 1:155197995:A:G | 1 | 1 | 1:155066988:C:T | 0.95 | $1.28 \times 10^{-18}$ | META | *EFNA4* | 10 | synonymous | *EFNA4* |
| 1:155197995:A:G | 2 | 3 | 1:155197995:A:G | 0.5 | $2.42 \times 10^{-17}$ | META | *MUC1* | 2.1 | intron | *THBS3* |
| 1:155197995:A:G | 3 | 20 | 1:155278322:A:T | 0.68 | $9.60 \times 10^{-6}$ | META | *FDPS* | 5.2 | intron | *HCN3* |
| 1:155197995:A:G | 4 | 4 | 1:155175305:G:A | 0.48 | $7.46 \times 10^{-10}$ | META | *MUC1* | 8.3 | upstream | *KRTCAP2* |
| 2:60480453:A:G | 1 | 7 | 2:60480759:G:A | 0.71 | $2.75 \times 10^{-9}$ | META | *BCL11A* | 7.4 | intron | *BCL11A* |
| 3:45818159:G:A | 1 | 5 | 3:45859597:C:T | 0.83 | $4.70 \times 10^{-204}$ | META | *CXCR6* | 0.14 | intron | - |
| 3:45818159:G:A | 2 | 1 | 3:45329214:G:A | 1 | $1.36 \times 10^{-62}$ | EUR | *LARS2* | 0.34 | - | - |
| 3:45818159:G:A | 3 | 1 | 3:45796521:G:T | 1 | $4.69 \times 10^{-18}$ | EUR | *FYCO1* | 9.2 | 5'UTR | *SLC6A20* |
| 3:45818159:G:A | 4 | 1 | 3:45239823:G:A | 1 | $4.07 \times 10^{-1}$ | SAS | *TMEM158* | 0.68 | - | - |
| 3:45818159:G:A | 5 | 13 | 3:46360250:G:A | 0.1 | $1.35 \times 10^{-1}$ | AFR | *CCR1* | 4.7 | 3'UTR | *CCR2* |
| 3:45818159:G:A | 6 | 1 | 3:45280485:C:T | 1 | $5.03 \times 10^{-1}$ | EUR | *TMEM158* | 7.6 | - | - |
| 3:45818159:G:A | 7 | 2 | 3:45645361:AAC:A | 0.66 | $8.56 \times 10^{-2}$ | SAS | *SACM1L* | 0.66 | intron | *LIMD1* |
| 3:45818159:G:A | 8 | 2 | 3:45570834:G:C | 0.5 | $4.27 \times 10^{-1}$ | SAS | *LIMD1* | 11 | - | - |
| 3:45818159:G:A | 9 | 2 | 3:45592155:A:T | 0.52 | $2.27 \times 10^{-1}$ | AFR | *LIMD1* | 1.5 | upstream | *LIMD1* |
| 3:146517122:G:A | 1 | 9 | 3:146517122:G:A | 0.17 | $2.28 \times 10^{-10}$ | EUR | *PLSCR1* | 23 | missense | *PLSCR1* |
| 3:169081115:G:GGAT | 1 | 36 | 3:169081067:C:T | 0.061 | $1.44 \times 10^{-9}$ | META | *MECOM* | 0.35 | downstream | *MECOM* |
| 3:169081115:G:GGAT | 2 | 9 | 3:169521790:A:G | 0.16 | $1.28 \times 10^{-6}$ | EUR | *MECOM* | 1.9 | intron | *MECOM* |
| 3:169081115:G:GGAT | 3 | 6 | 3:169299683:T:C | 0.54 | $1.30 \times 10^{-6}$ | EUR | *MECOM* | 4 | intron | *MECOM* |
| 3:169081115:G:GGAT | 4 | 1 | 3:169515899:A:AAATT | 1 | $2.32 \times 10^{-7}$ | META | *MECOM* | 0.67 | intron | *MECOM* |
| 4:105897896:G:A | 1 | 1 | 4:105897896:G:A | 1 | $4.65 \times 10^{-10}$ | EUR | *NPNT* | 24 | spliceregion | *NPNT* |
| 4:105897896:G:A | 2 | 1 | 4:105554756:C:CAT | 1 | $5.78 \times 10^{-8}$ | AFR | *PPA2* | 0.74 | intron | *ARHGEF38* |
| 5:132422622:A:G | 1 | 22 | 5:131995059:C:T | 0.47 | $7.23 \times 10^{-10}$ | EUR | *ACSL6* | 0.21 | intron | *ACSL6* |
| 7:100032719:C:T | 1 | 4 | 7:100032719:C:T | 0.5 | $8.43 \times 10^{-9}$ | META | *TRIM4* | 1.8 | intron | *ZKSCAN1* |
| 9:21206606:C:G | 1 | 3 | 9:21206606:C:G | 0.73 | $1.11 \times 10^{-11}$ | EUR | *IFNA10* | 24 | missense | *IFNA10* |
| 9:21206606:C:G | 2 | 2 | 9:21328701:G:A | 0.53 | $8.81 \times 10^{-9}$ | META | *KLHL9* | 11 | downstream | *KLHL9* |
| 9:133271182:T:C | 1 | 9 | 9:133271182:T:C | 0.84 | $2.82 \times 10^{-9}$ | META | - | 4.4 | intron | *ABO* |
| 11:34482745:G:A | 1 | 3 | 11:34482745:G:A | 0.58 | $5.13 \times 10^{-18}$ | META | *CAT* | 0.073 | intron | *ELF5* |
| 12:112919388:G:A | 1 | 60 | 12:112919388:G:A | 0.15 | $3.79 \times 10^{-10}$ | META | *OAS1* | 4.2 | spliceacceptor | *OAS1* |
| 12:132481571:G:A | 1 | 13 | 12:132479849:C:T | 0.14 | $1.34 \times 10^{-13}$ | META | *FBRSL1* | 0.94 | - | - |
| 12:132481571:G:A | 2 | 1 | 12:132158547:G:A | 1 | $3.86 \times 10^{-12}$ | EUR | *DDX51* | 0.31 | - | - |
| 13:112881427:C:T | 1 | 3 | 13:112886111:C:T | 0.4 | $3.31 \times 10^{-17}$ | META | *ATP11A* | 5.5 | 3'UTR | *ATP11A* |
| 15:31319838:G:T | 1 | 1 | 15:31334071:C:T | 0.99 | $1.34 \times 10^{-8}$ | EUR | *KLF13* | 5.6 | intron | *KLF13* |
| 15:31319838:G:T | 2 | 9 | 15:31319838:G:T | 0.22 | $1.48 \times 10^{-8}$ | META | *KLF13* | 2.7 | - | - |
| 16:29843264:G:A | 1 | 2 | 16:29843264:G:A | 0.93 | $1.84 \times 10^{-8}$ | AFR | *MVP* | 0.46 | intron | *MVP* |
| 16:29843264:G:A | 2 | 12 | 16:29709545:G:A | 0.11 | $4.16 \times 10^{-6}$ | EUR | *MVP* | 1.1 | downstream | - |
| 16:89196249:G:A | 1 | 2 | 16:89196249:G:A | 0.72 | $4.67 \times 10^{-10}$ | EUR | *SLC22A31* | 23 | missense | *SLC22A31* |
| 17:40003082:T:C | 1 | 66 | 17:40003082:T:C | 0.72 | $1.41 \times 10^{-9}$ | META | *MED24* | 2.2 | - | - |
| 17:49863260:C:A | 1 | 3 | 17:49863260:C:A | 0.75 | $9.48 \times 10^{-13}$ | META | *DLX3* | 5.4 | - | - |
| 17:49863260:C:A | 2 | 1 | 17:50213197:T:C | 1 | $1.41 \times 10^{-9}$ | META | *TMEM92* | 2.7 | intron | - |
| 17:49863260:C:A | 3 | 2 | 17:50212932:G:A | 0.88 | $4.33 \times 10^{-1}$ | SAS | *TMEM92* | 1.6 | intron | - |
| 17:49863260:C:A | 4 | 1 | 17:50030360:G:A | 0.96 | $1.58 \times 10^{-7}$ | EUR | *ITGA3* | 0.29 | - | - |
| 17:49863260:C:A | 5 | 2 | 17:50219800:G:A | 0.19 | $4.71 \times 10^{-1}$ | SAS | *TMEM92* | 12 | downstream | - |
| 17:49863260:C:A | 6 | 1 | 17:50109066:C:T | 1 | $3.11 \times 10^{-1}$ | EAS | *ITGA3* | 2.5 | downstream | *SAMD14* |
| 19:4717660:A:G | 1 | 1 | 19:4717660:A:G | 1 | $3.04 \times 10^{-51}$ | META | *DPP9* | 16 | intron | *DPP9* |
| 19:4717660:A:G | 2 | 1 | 19:5066102:C:T | 1 | $1.87 \times 10^{-14}$ | EUR | *KDM4B* | 0.2 | intron | *KDM4B* |
| 19:10352442:G:C | 1 | 1 | 19:10352442:G:C | 0.97 | $1.75 \times 10^{-26}$ | META | *TYK2* | 25 | missense | *TYK2* |
| 19:10352442:G:C | 2 | 1 | 19:10414696:G:A | 1 | $1.70 \times 10^{-11}$ | META | *ICAM1* | 1.1 | - | - |
| 19:48702888:G:C | 1 | 1 | 19:48496300:TC:T | 1 | $5.66 \times 10^{-15}$ | META | *LMTK3* | 0.78 | intron | *LMTK3* |
| 19:48702888:G:C | 2 | 10 | 19:48697960:C:T | 0.42 | $5.18 \times 10^{-13}$ | META | *RASIP1* | 2.4 | intron | *FUT2* |
| 19:48702888:G:C | 3 | 1 | 19:48500758:G:A | 1 | $1.14 \times 10^{-1}$ | META | *LMTK3* | 8 | intron | *LMTK3* |
| 19:48702888:G:C | 4 | 2 | 19:48494890:A:G | 0.48 | $3.32 \times 10^{-2}$ | META | *LMTK3* | 0.78 | intron | *LMTK3* |
| 21:33240996:A:G | 1 | 1 | 21:33412149:C:T | 1 | $1.53 \times 10^{-37}$ | EUR | *IFNAR2* | 2.2 | intron | *IFNGR2* |
| 21:33240996:A:G | 2 | 1 | 21:34355721:GGAAA:G | 1 | $1.29 \times 10^{-33}$ | EUR | *ATP5PO* | 0.63 | intron | - |
| 21:33240996:A:G | 3 | 9 | 21:33262748:G:GT | 0.73 | $8.91 \times 10^{-36}$ | META | *IFNAR2* | 0.85 | upstream | *IL10RB* |
| 21:33240996:A:G | 4 | 11 | 21:33954962:A:G | 0.19 | $4.58 \times 10^{-18}$ | META | *MRPS6* | 3.8 | intron | *LINC00649* |
| 21:33240996:A:G | 5 | 1 | 21:33441927:G:A | 1 | $1.97 \times 10^{-2}$ | EUR | *IFNAR2* | 0.48 | downstream | *IFNGR2* |
| 21:33240996:A:G | 6 | 30 | 21:33287696:C:T | 0.1 | $2.32 \times 10^{-7}$ | EUR | *IL10RB* | 1.4 | intron | *IL10RB* |
| 21:41471061:G:A | 1 | 24 | 21:41481267:TCAGACA:T | 0.28 | $4.07 \times 10^{-8}$ | META | *MX1* | 0.73 | intron | *TMPRSS2* |
| X:15602217:T:C | 1 | 1 | X:15752522:A:G | 1 | $6.95 \times 10^{-12}$ | EUR | *CA5B* | 2.6 | intron | *CA5B* |
| X:15602217:T:C | 2 | 1 | X:15602217:T:C | 1 | $5.10 \times 10^{-14}$ | META | *ACE2* | 8.6 | upstream | *ACE2* |
| X:15602217:T:C | 3 | 1 | X:15644354:C:T | 1 | $5.08 \times 10^{-9}$ | EUR | *CLTRN* | 2.5 | downstream | - |
| X:15602217:T:C | 4 | 14 | X:15616792:T:G | 0.12 | $9.91 \times 10^{-8}$ | AFR | *ACE2* | 3 | intron | *GS1-594A7.3* |

Table 5: Lead variants in credible sets from **multi-ancestry** fine-mapping in GenOMICC severe versus ctrl-all cohort. Locus index variant indicates index variant for locus as identified by the multi-study meta-analysis and is the variant with the highest posterior inclusion probability (PIP) in each credible set. *iCS* and *nCS* indicate the index and the number of variants in each credible set. *PIP* is the posterior inclusion probability of the lead variant. *MinP* indicates the minimum $P$-value for the lead variant among AFR, EAS, EUR, SAS or META from GenOMICC per population GWASs and multi-ancestry meta-analysis, while *MinP pop* indicates the analysis that produced the lowest $P$-value. *V2G gene* is the gene prioritised with the highest score by the Variant-to-Gene (V2G) pipeline on the Open Targets Genetics platform for the lead variant. *CADD* is the Combined Annotation Dependent Depletion score for the lead variant while *VEP consq* and *VEP gene* indicate the VEP predicted consequence on the canonical transcript and impacted gene, respectively.

| Locus index variant | iCS | nCS | CS worst variant | PIP | MinP | MinP pop | CADD | VEP consq | VEP gene |
|---|---|---|---|---|---|---|---|---|---|
| 1:155197995:A:G | 2 | 3 | 1:155202934:T:C | 0.33 | $6.10 \times 10^{-17}$ | META | 21 | missense | *THBS3* |
| 1:155197995:A:G | 3 | 20 | 1:155260340:C:T | 0.005 | $2.85 \times 10^{-4}$ | META | 2.5 | synonymous | *SCAMP3* |
| 17:40003082:T:C | 1 | 66 | 17:39974934:C:A | 0.0023 | $1.27 \times 10^{-6}$ | EUR | 22 | missense | *GSDMA* |
| 19:48702888:G:C | 2 | 10 | 19:48703346:C:T | 0.1 | $1.43 \times 10^{-12}$ | META | 7 | synonymous | *FUT2* |
| 21:41471061:G:A | 1 | 24 | 21:41473456:A:G | 0.0036 | $1.42 \times 10^{-6}$ | META | 1.8 | synonymous | *TMPRSS2* |

Table 6: Variants with worst consequence in credible sets from **multi-ancestry** fine-mapping in the GenOMICC severe versus ctrl-all cohort. Variants with the worst VEP consequence in each fine-mapped credible are shown, for cases when the worst variant is not the lead variant and the consequence has low impact or worse ("LOW", "MODERATE" or "HIGH" in VEP impact scale).*PIP*, *MinP*, *MinP pop*, *CADD*, *VEP consq* and *VEP gene* columns refer to the CS worst variant. *MinP* indicates the minimum *P*-value among AFR, EAS, EUR, SAS or META from GenOMICC per population GWASs and multi-ancestry meta-analysis, while *MinP pop* indicates the analysis that produced the lowest *P*-value. *CADD* is the Combined Annotation Dependent Depletion score while *VEP consq* and *VEP gene* indicate the VEP predicted consequence on the canonical transcript and impacted gene, respectively.

| Locus index variant | iCS | nCS | CS lead variant | PIP | P | V2G gene | CADD | VEP consq | VEP gene |
|---|---|---|---|---|---|---|---|---|---|
| 1:64951326:A:G | 1 | 4 | 1:64951326:A:G | 0.94 | $2.16 \times 10^{-9}$ | *JAK1* | 2.3 | intron | *JAK1* |
| 1:155197995:A:G | 1 | 1 | 1:155036692:C:A | 0.97 | $1.74 \times 10^{-9}$ | *DCST1* | 3.3 | intron | *DCST1* |
| 1:155197995:A:G | 2 | 3 | 1:155197995:A:G | 0.56 | $1.41 \times 10^{-16}$ | *MUC1* | 2.1 | intron | *THBS3* |
| 1:155197995:A:G | 3 | 2 | 1:155066988:C:T | 0.86 | $6.58 \times 10^{-17}$ | *EFNA4* | 10 | synonymous | *EFNA4* |
| 1:155197995:A:G | 4 | 33 | 1:155278322:A:T | 0.45 | $4.58 \times 10^{-4}$ | *FDPS* | 5.2 | intron | *HCN3* |
| 1:155197995:A:G | 5 | 5 | 1:155302683:CT:C | 0.45 | $6.12 \times 10^{-8}$ | *RUSC1* | 0.72 | upstream | *PKLR* |
| 2:60480453:A:G | 1 | 11 | 2:60480453:A:G | 0.18 | $1.86 \times 10^{-7}$ | *BCL11A* | 10 | intron | *BCL11A* |
| 3:45818159:G:A | 1 | 1 | 3:45329214:G:A | 1 | $1.36 \times 10^{-62}$ | *LARS2* | 0.34 | - | - |
| 3:45818159:G:A | 2 | 1 | 3:45796521:G:T | 1 | $4.69 \times 10^{-18}$ | *FYCO1* | 9.2 | 5'UTR | *SLC6A20* |
| 3:45818159:G:A | 3 | 1 | 3:45239823:G:A | 1 | $7.38 \times 10^{-1}$ | *TMEM158* | 0.68 | - | - |
| 3:45818159:G:A | 4 | 1 | 3:45280485:C:T | 1 | $5.03 \times 10^{-1}$ | *TMEM158* | 7.6 | - | - |
| 3:45818159:G:A | 5 | 13 | 3:46193413:A:T | 0.077 | $6.71 \times 10^{-1}$ | *CCR3* | 1.7 | - | - |
| 3:45818159:G:A | 6 | 2 | 3:45696232:C:T | 0.5 | $4.65 \times 10^{-1}$ | *SACM1L* | 1.9 | intron | *SACM1L* |
| 3:45818159:G:A | 7 | 2 | 3:45569212:T:C | 0.5 | $9.21 \times 10^{-1}$ | *LIMD1* | 5.2 | - | - |
| 3:45818159:G:A | 8 | 2 | 3:45570834:G:C | 0.5 | $6.92 \times 10^{-1}$ | *LIMD1* | 11 | - | - |
| 3:45818159:G:A | 9 | 7 | 3:45859597:C:T | 0.74 | $5.90 \times 10^{-178}$ | *CXCR6* | 0.14 | intron | - |
| 3:146517122:G:A | 1 | 9 | 3:146517122:G:A | 0.18 | $2.28 \times 10^{-10}$ | *PLSCR1* | 23 | missense | *PLSCR1* |
| 3:169081115:G:GGAT | 1 | 1 | 3:169515899:A:AAATT | 1 | $6.20 \times 10^{-7}$ | *MECOM* | 0.67 | intron | *MECOM* |
| 3:169081115:G:GGAT | 2 | 14 | 3:169521598:C:T | 0.15 | $1.98 \times 10^{-1}$ | *MECOM* | 2.4 | intron | *MECOM* |
| 3:169081115:G:GGAT | 3 | 29 | 3:169523892:T:TACAC | 0.12 | $7.12 \times 10^{-8}$ | *MECOM* | 0.68 | intron | *MECOM* |
| 4:105897896:G:A | 1 | 1 | 4:105897896:G:A | 1 | $4.65 \times 10^{-10}$ | *NPNT* | 24 | spliceregion | *NPNT* |
| 6:41520640:G:A | 1 | 8 | 6:41522116:T:C | 0.25 | $1.45 \times 10^{-7}$ | *FOXP4* | 5.2 | downstream | *LINC01276* |
| 7:100032719:C:T | 1 | 4 | 7:100032719:C:T | 0.48 | $7.96 \times 10^{-8}$ | *TRIM4* | 1.8 | intron | *ZKSCAN1* |
| 9:21206606:C:G | 1 | 2 | 9:21328701:G:A | 0.53 | $3.38 \times 10^{-8}$ | *KLHL9* | 11 | downstream | *KLHL9* |
| 9:21206606:C:G | 2 | 3 | 9:21206606:C:G | 0.77 | $1.11 \times 10^{-11}$ | *IFNA10* | 24 | missense | *IFNA10* |
| 9:133271182:T:C | 1 | 11 | 9:133271182:T:C | 0.79 | $1.32 \times 10^{-8}$ | - | 4.4 | intron | *ABO* |
| 11:34482745:G:A | 1 | 3 | 11:34482745:G:A | 0.58 | $1.62 \times 10^{-14}$ | *CAT* | 0.073 | intron | *ELF5* |
| 12:112919388:G:A | 1 | 64 | 12:112919388:G:A | 0.11 | $5.02 \times 10^{-9}$ | *OAS1* | 4.2 | spliceacceptor | *OAS1* |
| 12:132481571:G:A | 1 | 1 | 12:132158547:G:A | 1 | $3.86 \times 10^{-12}$ | *DDX51* | 0.31 | - | - |
| 12:132481571:G:A | 2 | 12 | 12:132537984:G:A | 0.19 | $2.70 \times 10^{-11}$ | *FBRSL1* | 2.9 | intron | *FBRSL1* |
| 12:132481571:G:A | 3 | 12 | 12:132536375:T:C | 0.28 | $1.76 \times 10^{-3}$ | *POLE* | 4 | intron | *FBRSL1* |
| 13:112881427:C:T | 1 | 4 | 13:112882313:A:G | 0.31 | $1.49 \times 10^{-13}$ | *ATP11A* | 0.1 | 3'UTR | *ATP11A* |
| 15:31319838:G:T | 1 | 1 | 15:31334071:C:T | 1 | $1.34 \times 10^{-8}$ | *KLF13* | 5.6 | intron | *KLF13* |
| 16:29843264:G:A | 1 | 12 | 16:29881840:G:A | 0.08 | $4.16 \times 10^{-6}$ | *SEZ6L2* | 2 | intron | *SEZ6L2* |
| 16:89196249:G:A | 1 | 2 | 16:89196249:G:A | 0.74 | $4.67 \times 10^{-10}$ | *SLC22A31* | 23 | missense | *SLC22A31* |
| 17:49863260:C:A | 1 | 1 | 17:50213197:T:C | 1 | $2.16 \times 10^{-9}$ | *TMEM92* | 2.7 | intron | - |
| 17:49863260:C:A | 2 | 1 | 17:50030360:G:A | 0.99 | $1.58 \times 10^{-7}$ | *ITGA3* | 0.29 | - | - |
| 17:49863260:C:A | 3 | 1 | 17:50109066:C:T | 1 | $6.74 \times 10^{-1}$ | *ITGA3* | 2.5 | downstream | *SAMD14* |
| 17:49863260:C:A | 4 | 2 | 17:50212932:G:A | 0.94 | $5.09 \times 10^{-1}$ | *TMEM92* | 1.6 | intron | - |
| 17:49863260:C:A | 5 | 2 | 17:50225230:G:T | 0.7 | $7.19 \times 10^{-1}$ | *COL1A1* | 1 | - | - |
| 17:49863260:C:A | 6 | 3 | 17:49863260:C:A | 0.69 | $3.85 \times 10^{-11}$ | *DLX3* | 5.4 | - | - |
| 19:4717660:A:G | 1 | 1 | 19:4717660:A:G | 0.99 | $1.40 \times 10^{-46}$ | *DPP9* | 16 | intron | *DPP9* |
| 19:4717660:A:G | 2 | 1 | 19:5066102:C:T | 1 | $1.87 \times 10^{-14}$ | *KDM4B* | 0.2 | intron | *KDM4B* |
| 19:10352442:G:C | 1 | 1 | 19:10352442:G:C | 0.98 | $2.31 \times 10^{-25}$ | *TYK2* | 25 | missense | *TYK2* |
| 19:10352442:G:C | 2 | 1 | 19:10414696:G:A | 1 | $1.85 \times 10^{-11}$ | *ICAM1* | 1.1 | - | - |
| 19:10352442:G:C | 3 | 1 | 19:10020130:A:C | 0.95 | $8.14 \times 10^{-7}$ | *RDH8* | 2.8 | intron | *RDH8* |
| 19:48702888:G:C | 1 | 1 | 19:48496300:TC:T | 1 | $9.88 \times 10^{-14}$ | *LMTK3* | 0.78 | intron | *LMTK3* |
| 19:48702888:G:C | 2 | 1 | 19:48500758:G:A | 1 | $2.31 \times 10^{-1}$ | *LMTK3* | 8 | intron | *LMTK3* |
| 19:48702888:G:C | 3 | 5 | 19:48466475:C:T | 0.25 | $6.18 \times 10^{-1}$ | *CYTH2* | 0.14 | 3'UTR | *KCNJ14* |
| 19:48702888:G:C | 4 | 9 | 19:48697960:C:T | 0.52 | $5.23 \times 10^{-13}$ | *RASIP1* | 2.4 | intron | *FUT2* |
| 21:33240996:A:G | 1 | 1 | 21:33412149:C:T | 1 | $1.53 \times 10^{-37}$ | *IFNAR2* | 2.2 | intron | *IFNGR2* |
| 21:33240996:A:G | 2 | 1 | 21:34355721:GGAAA:G | 1 | $1.29 \times 10^{-33}$ | *ATP5PO* | 0.63 | intron | - |
| 21:33240996:A:G | 3 | 1 | 21:33441927:G:A | 1 | $1.97 \times 10^{-2}$ | *IFNAR2* | 0.48 | downstream | *IFNGR2* |
| 21:33240996:A:G | 4 | 18 | 21:33232252:G:A | 0.094 | $1.59 \times 10^{-25}$ | *IL10RB* | 0.43 | intron | *IFNAR2* |
| 21:33240996:A:G | 5 | 14 | 21:33954962:A:G | 0.15 | $8.39 \times 10^{-15}$ | *MRPS6* | 3.8 | intron | *LINC00649* |
| 21:33240996:A:G | 6 | 31 | 21:33287378:C:T | 0.097 | $6.15 \times 10^{-11}$ | *IFNAR1* | 3.6 | intron | *IL10RB* |
| X:15602217:T:C | 1 | 1 | X:15752522:A:G | 1 | $6.95 \times 10^{-12}$ | *CA5B* | 2.6 | intron | *CA5B* |
| X:15602217:T:C | 2 | 1 | X:15644354:C:T | 1 | $5.08 \times 10^{-9}$ | *CLTRN* | 2.5 | downstream | - |

Table 7: Lead variants in credible sets from **European (EUR)** fine-mapping in the GenOMICC severe versus ctrl-all cohort. Locus index variant indicates index variant for locus as identified by the multi-study meta-analysis and is the variant with the highest posterior inclusion probability (PIP) in each credible set. *iCS* and *nCS* indicate the index and the number of variants in each credible set. *PIP* is the posterior inclusion probability of the lead variant. *P* indicates the *P*-value in the EUR GWAS for the lead variant. *V2G gene* is the gene prioritised with the highest score by the Variant-to-Gene (V2G) pipeline on the Open Targets Genetics platform for the lead variant. *CADD* is the Combined Annotation Dependent Depletion score for the lead variant while *VEP consq* and *VEP gene* indicate the VEP predicted consequence on the canonical transcript and impacted gene, respectively.

| Locus index variant | iCS | nCS | CS worst variant | PIP | P | CADD | VEP consq | VEP gene |
|---|---|---|---|---|---|---|---|---|
| 1:155197995:A:G | 2 | 3 | 1:155202934:T:C | 0.29 | $5.34 \times 10^{-16}$ | 21 | missense | *THBS3* |
| 1:155197995:A:G | 4 | 33 | 1:155260340:C:T | 0.009 | $8.14 \times 10^{-3}$ | 2.5 | synonymous | *SCAMP3* |
| 9:133271182:T:C | 1 | 11 | 9:133257521:T:TC | 0.0072 | $2.00 \times 10^{-6}$ | 15 | frameshift | *ABO* |
| 19:48702888:G:C | 4 | 9 | 19:48703346:C:T | 0.1 | $3.41 \times 10^{-12}$ | 7 | synonymous | *FUT2* |
| 21:33240996:A:G | 4 | 18 | 21:33241950:T:G | 0.057 | $2.66 \times 10^{-25}$ | 0.002 | missense | *IFNAR2* |

Table 8: Variants with worst consequence in credible sets from **European (EUR)** fine-mapping in the GenOMICC severe versus ctrl-all cohort. Variants with the worst VEP consequence in each fine-mapped credible are shown, for cases when the worst variant is not the lead variant in terms of PIP in the credible set and the consequence has low impact or worse ("LOW", "MODERATE" or "HIGH" in VEP impact scale). *PIP*, *P*, *CADD*, *VEP consq* and *VEP gene* columns refer to the CS worst variant. *P* indicates *P*-values in the EUR GWAS. *CADD* is the Combined Annotation Dependent Depletion score while *VEP consq* and *VEP gene* indicate the VEP predicted consequence on the canonical transcript and impacted gene, respectively.

###### 1.1.4 Age and Sex Stratified analysis

We performed an age-stratified analysis for European ancestry individuals dividing them between $< 60$ and $>= 60$ years old (the median age for cases; Table 1). We also performed a sex-stratified analysis for European ancestry individuals. in both cases, we compared the two cohorts using the same methodology as[12]. We did not find genome wide significant differences in effects for either analysis. In Fig. 5 we show a comparison of the effects at the lead variants of our main analysis.

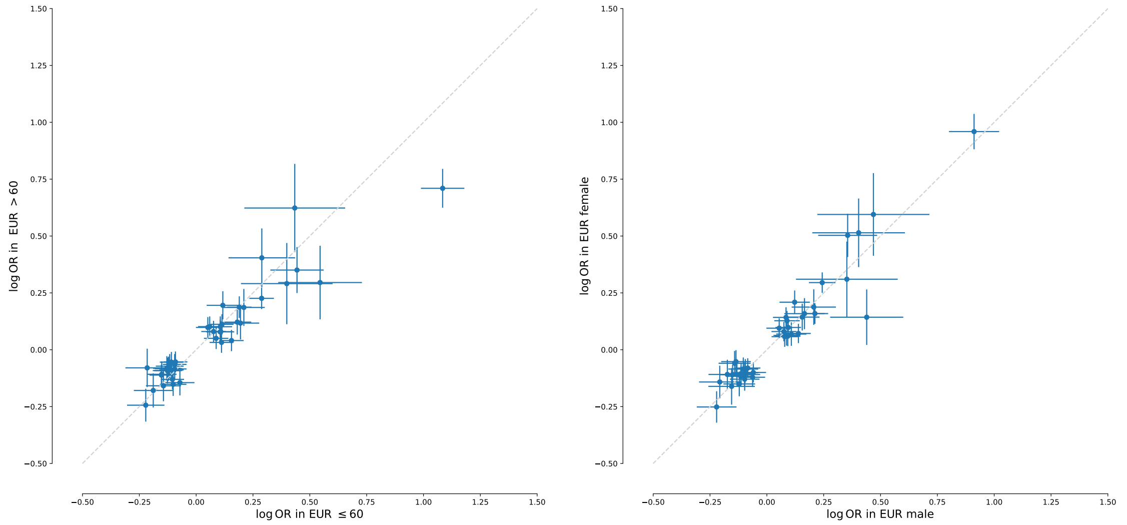

Figure 5: Comparison of logOR estimates with 95% CI at lead variants of the main analysis for age and sex stratified GWAS in European ancestry individuals.

##### 1.2 Polygenic risk score (PRS) analysis

With 80% of the critical cases vs. ctrl-mld cohort we calculated a polygenic risk score (PRS) for individuals of European ancestry. We then tested the PRS on the remaining 20% of individuals from European ancestry. We compared the genetic PRS to a risk score based on covariates comprising sex and age. A model including PRS and covariates improved prediction of Covid-19 cases (auc=0.73) compared with a model using just covariates (auc=0.70). The mean age of the top 5% individuals with critical Covid-19 is 58.83 and 62% of them are males compared to a mean age of 60.1 and a percentage of 66% of males in our test dataset.

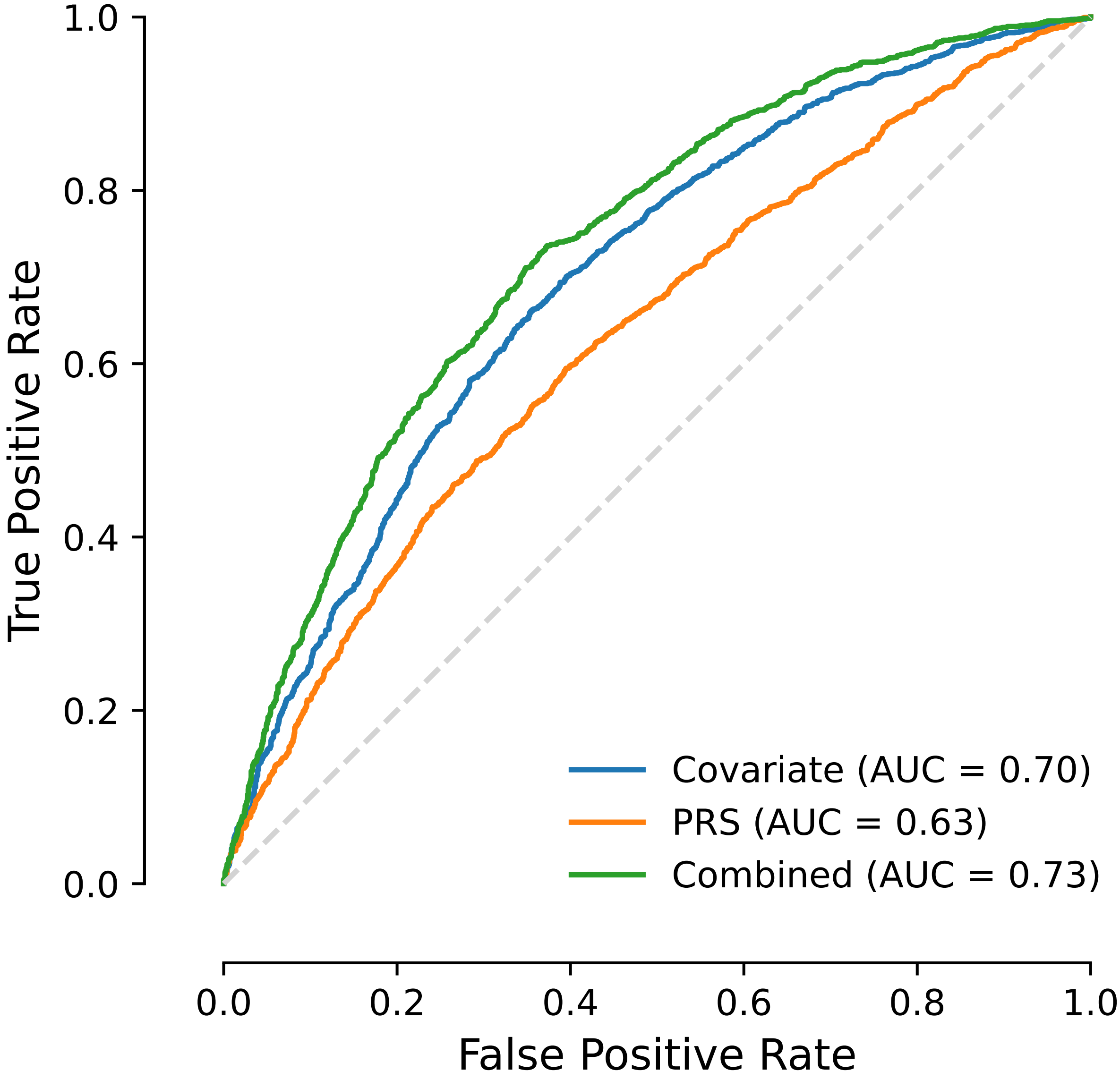

Figure 6: Receiver operator curves for risk scores on validation cohort of European ancestry, comparing a covariate (Covariate) and genetic (PRS) risk scores, as well as their sum (Combined).

#### 1.3 Gene-based rare variant analysis tests (RVAT)

##### 1.3.1 RVAT Cohorts

For the gene-based rare variant association analysis, we used a subset of 28,091 individuals from our cohorts, consisting of 11,423 individuals with critical COVID-19 from GenOMICC and 16,668 control individuals. The control group included 11,268 individuals from the mild COVID-19 cohort and 5,400 from the 100kGP cohort, all processed with the same alignment and variant calling pipeline (a control set referred to as ctrl-dgn; Extended Data Fig.1; Methods). The reasoning for using the ctrl-dgn cohort for the RVAT analysis is as follows: (1) We used a subset of controls processed with the same pipeline as opposed to using all available controls (ctrl-all) to minimise potential batch effects as those would be more difficult to quality control than our GWAS analysis on common variants. (2) We included 100kGP individuals in the controls to increase discovery power, boosting control numbers by ∼50% compared to using only ctrl-mld.

Six additional studies contributed data for our multi-study RVAT meta-analysis. Case/control breakdowns across all contributing studies are shown in Extended Data Fig.1 and Table 9.

| Study cohort | Ancestry | $N_{cases}$ | $N_{controls}$ |
|---|---|---|---|
| GenOMICC | EUR | 9,128 | 14,670 |
| GenOMICC | SAS | 1,229 | 1,165 |
| GenOMICC | AFR | 681 | 594 |
| GenOMICC | EAS | 385 | 239 |
| UK Biobank | EUR | 2,101 | 465,275 |
| GENCOVID | EUR | 1,362 | 705 |
| BQC19-SweCovid | EUR | 361 | 1,143 |
| DeCOI | EUR | 243 | 587 |
| POLCOVID | EUR | 109 | 1249 |
| PMBB | AFR | 107 | 10,264 |
| PMBB | EUR | 80 | 28,964 |
| Total | - | 15,786 | 524,855 |

Table 9: Sample sizes for each study cohort included in the multi-study RVAT meta-analysis.

##### 1.3.2 Multi-study RVAT meta-analysis

To assess the role of rare variants in critical illness, we conducted gene-based analysis with REGENIE[13]. We defined two sets of variant masks: *strict_LoF* including variants with predicted loss of function effects

(LoF) and *mild_LoF*, including variants annotated as either LoF or missense (Methods). To maximise power, we combined strength across multiple variant tests (Burden[14], SKAT-O[15], ACAT-V[16]) and allele frequency thresholds (singletons, AF=0.005) with the Cauchy combination test and calculated a single $P$-value (ACAT-O; Methods). We then meta-analysed gene-wise ACAT-O $P$-values across all studies (Table 9) using Stouffer's method and weighted by effective sample size ($n_w$; Methods). We set a Bonferroni corrected gene-wide significance threshold of $2.58\times10^{-6}$ (0.05 divided by 19,357 protein coding genes tested).

From the multi-study RVAT meta-analysis, we obtained *TLR7* as gene-wide significant for the *strict_LoF* mask ($P$-value=$8.25\times10^{-8}$; Figure 7A) and *SLC50A1* and *TLR7* as gene-wide significant for the *mild_LoF* mask ($P$-value $= 1.04\times10^{-6}$ and $1.74\times10^{-6}$, respectively; Figure 7B). When considering contributions from each study, *SLC50A1* was nominally significant ($P < 0.05$) in three studies (GenOMICC, UK Biobank and BQC19-SweCovid), while *TLR7* was nominally significant in two studies (GenOMICC and UK Biobank), Table 10.

To replicate the novel *SLC50A1* signal for the *mild_LoF* mask, we performed a leave-largest-study-out meta-analysis, excluding the GenOMICC cohorts and meta-analysing the remaining studies obtaining a nominally significant $P$-value ($P$=0.0289) for *SLC50A1*.

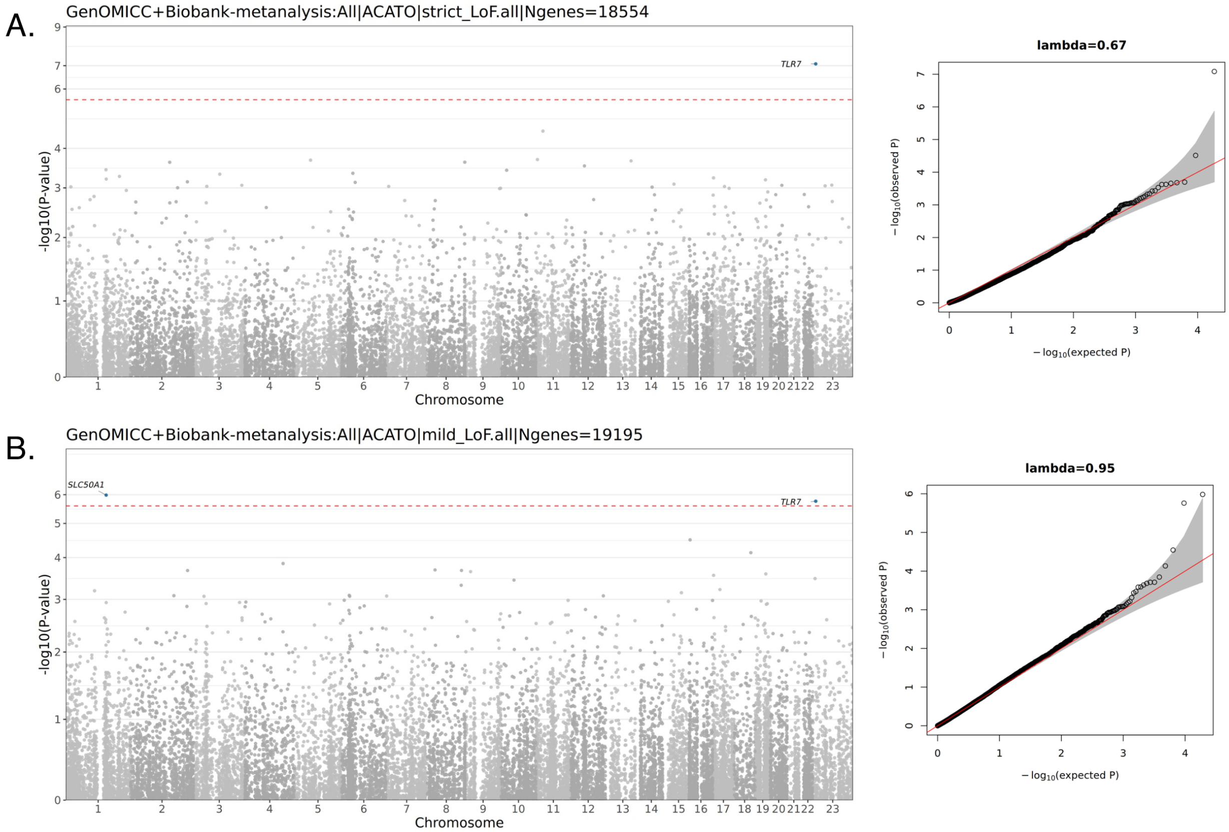

Figure 7: Results from rare variant multi-study meta-analysis. A. Results for *strict_lof* mask. B. Results for *mild_lof* mask. Genes that are gene-wide significant at Bonferroni corrected threshold $P < 2.58 \times 10^{-6}$ (red dashed line) are highlighted in blue. ACAT-O gene-based $P$-values are shown.

| Cohort | Gene | Mask | P | OR CI burden | Best test | Best test P |
|---|---|---|---|---|---|---|
| GenOMICC-EUR | *SLC50A1* | mild-LoF | $1.53 \times 10^{-5}$* | 1.91[1.42-2.56] | SKATO | $1.21 \times 10^{-5}$ |
| GenOMICC-EUR | *SLC50A1* | strict-LoF | $4.11 \times 10^{-1}$ | 2.1[0.359-12.2] | ACATV | $4.11 \times 10^{-1}$ |
| GenOMICC-EUR | *TLR7* | mild-LoF | $3.99 \times 10^{-7}$* | 1.77[1.36-2.3] | SKATO | $2.81 \times 10^{-7}$ |
| GenOMICC-EUR | *TLR7* | strict-LoF | $5.32 \times 10^{-6}$* | 8.7[3.43-22.1] | ACATV | $5.32 \times 10^{-6}$ |
| GenOMICC-SAS | *SLC50A1* | mild-LoF | $7.16 \times 10^{-2}$ | 12.6[0.801-199] | ACATV | $7.16 \times 10^{-2}$ |
| GenOMICC-SAS | *TLR7* | mild-LoF | $2.68 \times 10^{-1}$ | 1.48[0.741-2.94] | ACATV | $2.68 \times 10^{-1}$ |
| GenOMICC-SAS | *TLR7* | strict-LoF | $1.22 \times 10^{-1}$ | 3.27[0.729-14.7] | ACATV | $1.22 \times 10^{-1}$ |
| GenOMICC-AFR | *SLC50A1* | mild-LoF | $2.55 \times 10^{-1}$ | 5.31[0.299-94.3] | ACATV | $2.55 \times 10^{-1}$ |
| GenOMICC-AFR | *SLC50A1* | strict-LoF | $4.35 \times 10^{-1}$ | 5.04[0.0865-294] | ACATV | $4.35 \times 10^{-1}$ |
| GenOMICC-AFR | *TLR7* | mild-LoF | $2.42 \times 10^{-1}$ | 2.26[0.576-8.87] | ACATV | $2.42 \times 10^{-1}$ |
| GenOMICC-AFR | *TLR7* | strict-LoF | $5.73 \times 10^{-1}$ | 4.01[0.0319-505] | ACATV | $5.73 \times 10^{-1}$ |
| GenOMICC-EAS | *SLC50A1* | mild-LoF | $1.65 \times 10^{-1}$ | 0.0506[0.000746-3.43] | ACATV | $1.65 \times 10^{-1}$ |
| GenOMICC-EAS | *TLR7* | mild-LoF | $6.22 \times 10^{-1}$ | 3.62[0.0215-611] | ACATV | $6.22 \times 10^{-1}$ |
| DeCOI | *SLC50A1* | mild-LoF | $8.35 \times 10^{-1}$ | 0.838[0.159-4.43] | ACATV | $8.35 \times 10^{-1}$ |
| DeCOI | *TLR7* | mild-LoF | $3.18 \times 10^{-1}$ | 0.471[0.107-2.06] | ACATV | $3.18 \times 10^{-1}$ |
| GENCOVID | *SLC50A1* | mild-LoF | $6.52 \times 10^{-1}$ | 1.31[0.409-4.17] | ACATV | $6.52 \times 10^{-1}$ |
| GENCOVID | *TLR7* | mild-LoF | $4.47 \times 10^{-1}$ | 1.57[0.49-5.05] | ACATV | $4.47 \times 10^{-1}$ |
| PMBB-EUR | *SLC50A1* | mild-LoF | $2.57 \times 10^{-1}$ | 2.94[0.171-50.5] | ACATV | $2.45 \times 10^{-1}$ |
| PMBB-EUR | *SLC50A1* | strict-LoF | $9.60 \times 10^{-1}$ | 0.367[5.04e-18-2.67e+16] | ACATV | $9.60 \times 10^{-1}$ |
| PMBB-AFR | *SLC50A1* | mild-LoF | $6.99 \times 10^{-1}$ | 0.35[0.0175-7] | FIRTH | $4.92 \times 10^{-1}$ |
| UK Biobank | *SLC50A1* | mild-LoF | $2.81 \times 10^{-2}$* | 1.19[0.685-2.07] | SKATO | $2.56 \times 10^{-2}$ |
| UK Biobank | *SLC50A1* | strict-LoF | $5.51 \times 10^{-1}$ | 0.36[0.0381-3.4] | FIRTH | $3.73 \times 10^{-1}$ |
| UK Biobank | *TLR7* | mild-LoF | $4.21 \times 10^{-1}$ | 1.01[0.945-1.07] | ACATV | $2.09 \times 10^{-1}$ |
| UK Biobank | *TLR7* | strict-LoF | $1.50 \times 10^{-3}$* | 13.9[2.74-70.8] | ACATV | $1.50 \times 10^{-3}$ |
| BQC19-SweCovid | *SLC50A1* | mild-LoF | $4.05 \times 10^{-2}$* | 5.13[1.07-24.5] | ACATV | $4.05 \times 10^{-2}$ |
| BQC19-SweCovid | *SLC50A1* | strict-LoF | $6.52 \times 10^{-1}$ | 0.295[0.00146-59.4] | ACATV | $6.52 \times 10^{-1}$ |
| BQC19-SweCovid | *TLR7* | mild-LoF | $9.96 \times 10^{-1}$ | 0.995[0.17-5.83] | ACATV | $9.96 \times 10^{-1}$ |

Table 10: Results for significant genes for RVAT analysis across studies. $P$ indicates ACAT-O gene-based $P$-values with asterisk (*) indicating nominally significant $P$-value ($P < 0.05$) in the respective cohort and mask. *OR CI burden* shows the odds ratio and 95% Confidence interval (CI) for the burden test, *Best test* shows the test that produced the minimum $P$-value among the combined tests FIRTH (burden), SKATO, ACATV within ACAT-O test. *Best test P* shows the $P$-value of the displayed *best test.* Cohorts and mask combinations that yielded no variants in the respective genes after applying the respective mask are not shown.

##### 1.3.3 Individual variants contributing to the RVAT gene signals

We examined individual variant contributions from the EUR GenOMICC vs ctrl-dgn analysis to the *SLC50A1* and *TLR7* signals to understand the genetic architecture of the association signals by estimating their effect sizes with Firth's logistic regression as implemented in REGENIE. Gene *SLC50A1* had 24 missense and 3 LoF mutations. We identified *SLC50A1* missense variant 1:155138217:G:T with minor allele frequency (maf)=0.00242 ($P$-value = $2.03 \times 10^{-5}$, OR[95%] = 2.31[1.57-3.39], CADD=28.6) as the only contributing variant with $P$-value $< 0.05$. Gene *TLR7* had 11 LoF and 45 missense mutations and there were no variants with minor allele count (MAC) >= 10 with $P$-values $< 0.05$. Two *TLR7* variants with MAC $< 10$, LoF variant X:12885886:CACTT:C ($P$-value $= 1.9 \times 10^{-2}$, OR[95%] = 5.05[1.31-19.6], CADD=13.2) and missense variant X:12886145:G:C($P$-value $= 9.66 \times 10^{-3}$, OR[95%] = 7.43[1.63-34], CADD=14.5), were nominally significant ($P < 0.05$).

##### 1.3.4 Conditional analyses for *SLC50A1*

The *SLC50A1* gene signal was identified as gene-wide significant in the multi-study RVAT meta-analysis. The gene resides within a GWAS significant locus marked by the index variant 1:155197995:A:G (rs41264915) at 1q22 (Table 2). We conducted conditional RVAT analyses to assess whether the *SLC50A1* RVAT signal is conditionally independent of the common variant signals identified through cross-ancestry or EUR fine-mapping within that locus (Table 5, and Table 7, respectively). We used the GenOMICC severe vs ctrl-dgn EUR cohort results for these conditional analyses as it has the largest number of cases and is the greatest contributor to the RVAT signal. Across conditioned variants, the *SLC50A1* gene signal $P$-value was reduced in significance only when conditioning on the common variant 1:155036692:C:A that was fine-mapped in EUR (Table 11). Therefore, we hypothesise that lead variant signal 1:155036692:C:A is partially tagging the *SLC50A1* gene signal.

| CondVar | Source | Pval |
|---|---|---|
| 1:155066988:C:T | xancestry/EUR | $1.12 \times 10^{-5}$ |
| 1:155197995:A:G | xancestry/EUR | $1.08 \times 10^{-5}$ |
| 1:155278322:A:T | xancestry/EUR | $1.06 \times 10^{-5}$ |
| 1:155175305:G:A | xancestry | $1.35 \times 10^{-5}$ |
| 1:155036692:C:A | EUR | $2.79 \times 10^{-3}$ |
| 1:155302683:CT:C | EUR | $1.33 \times 10^{-5}$ |
| - | - | $1.53 \times 10^{-5}$ |

Table 11: Conditional analyses for the RVAT *SLC50A1* gene signal in EUR GenOMICC severe vs ctrl-dgn cohort for the *mild_lof* mask. *CondVar* is the conditioned lead variant from the credible sets identified in multi-ancestry fine-mapping (xancestry), EUR fine-mapping (EUR), or both (xancestry/EUR). Last row presents unconditional results. $P$-values shown (column: Pval) are from the ACAT-O gene-based test.

##### 1.3.5 RVAT analysis stratified by age

Te explore genetic effects for COVID-19 severity in younger versus older individuals, we conducted RVAT multi-ancestry meta-analysis on GenOMICC severe vs ctrl-dgn cohort subsets that were stratified by age. As age 60 was the median age in cases (Table 1), we split the cohorts in age<60 and age>=60 to evenly split the dataset and have comparable statistical power in the two groups. The age<60 analysis re-produced the signals for the *mild_LoF* mask for both *SLC50A1* and *TLR7* ($P$-value $= 1.11 \times 10^{-6}$ and $8.35 \times 10^{-8}$, respectively; Figure 8A). For age>=60 we observed attenuation of both the *SLC50A1* and *TLR7* signals

(Figure 8B). The *strict_LoF* mask produced a significant signal in the age<60 analysis for gene *VPS13B* ($P$-value $= 1.78 \times 10^{-6}$; Figure 9A) and none in the age>=60 cohort (Figure 9B). As *VPS13B* was not significant in the multi-study meta-analysis ($P$-values of 0.85 and 0.63 for the *strict_LoF* nor *mild_LoF* masks, respectively), we did not follow it up further.

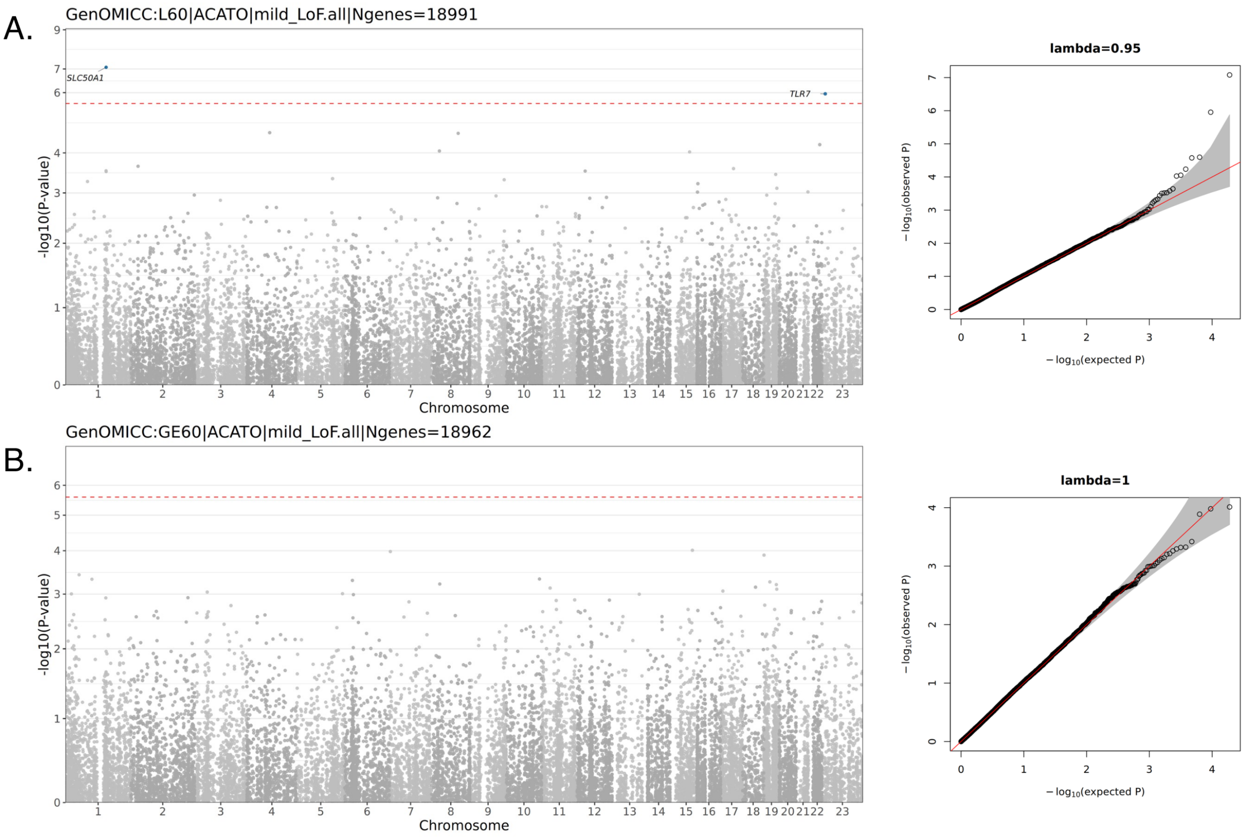

Figure 8: Results from rare variant multi-ancestry meta-analysis for cohorts stratified by age for the *mild_lof* mask. A. Results for individuals with age $< 60$; B. Results for individuals with age $>= 60$. Genes that are gene-wide significant at Bonferroni corrected threshold $P < 2.58 \times 10^{-6}$ (red dashed line) are highlighted in blue. ACAT-O gene-based $P$-values are shown.

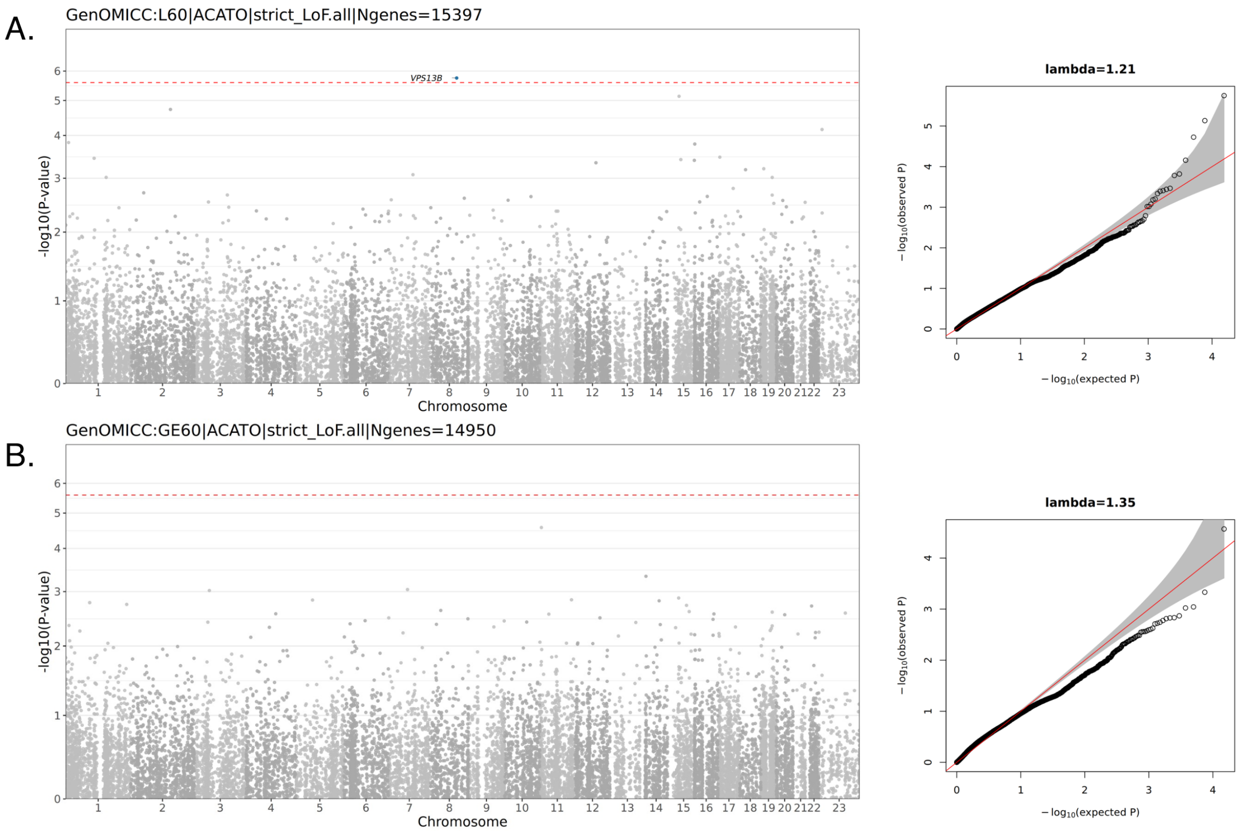

Figure 9: Results from rare variant multi-ancestry meta-analysis for cohorts stratified by age for the *strict_lof* mask. A. Results for individuals with age < 60; B. Results for individuals with age >= 60. Genes that are gene-wide significant at Bonferroni corrected threshold $P < 2.58 \times 10^{-6}$ (red dashed line) are highlighted in blue. ACAT-O gene-based $P$-values are shown.

### 2 Additional figures

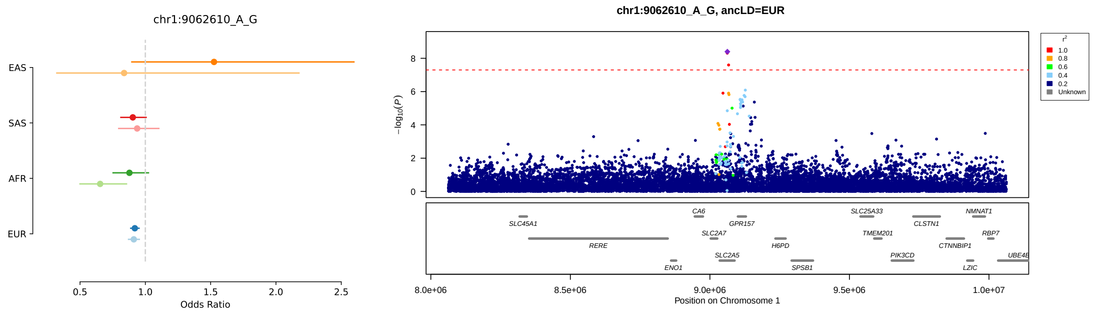

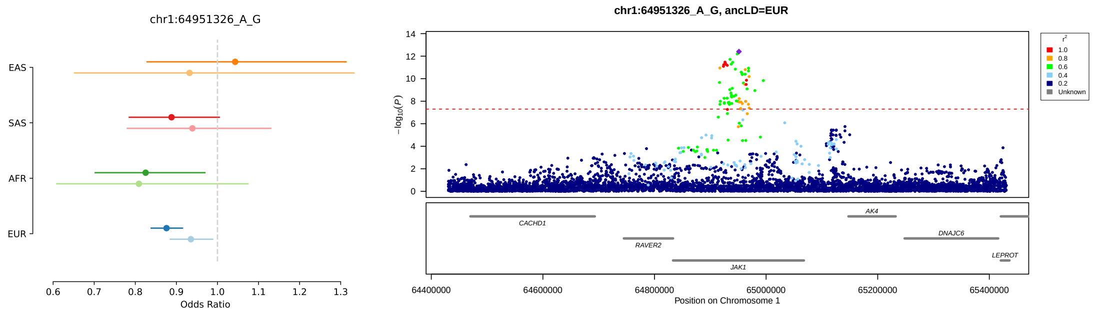

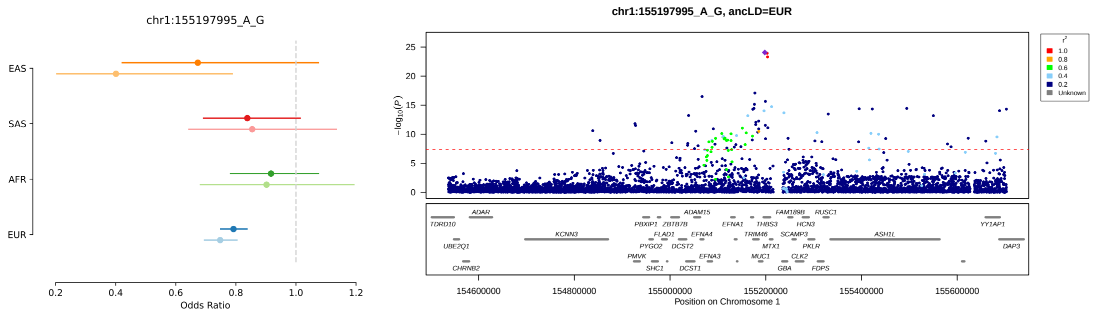

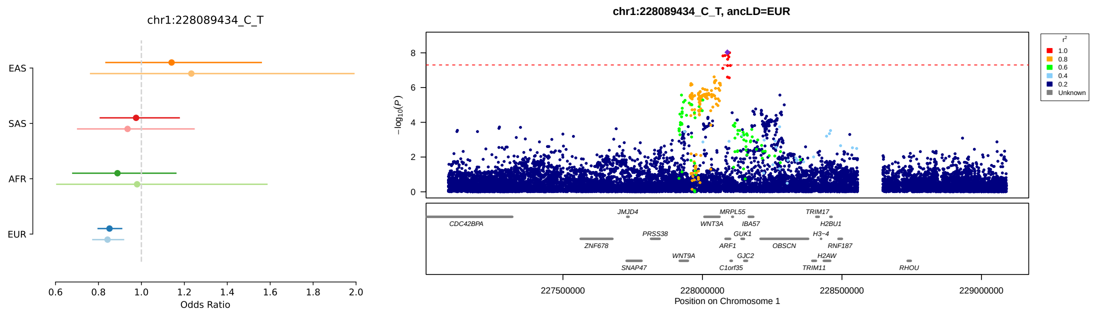

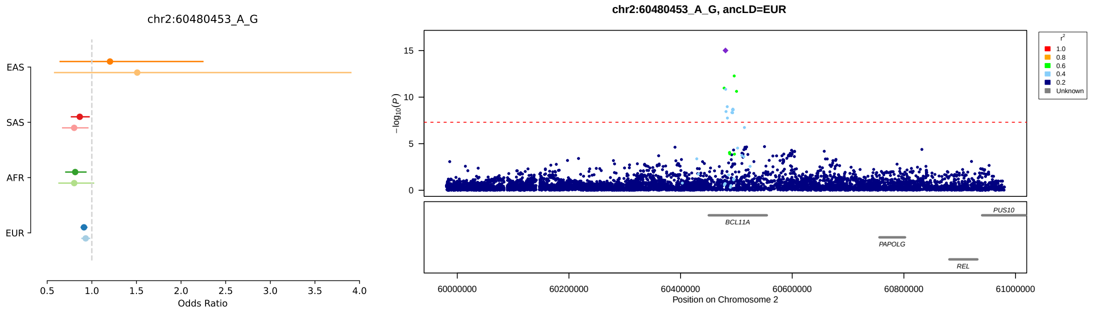
chr2:60480453_A_G
EAS
SAS
AFR
EUR
Odds Ratio
chr2:60480453_A_G, ancLD=EUR
−log10(P)
r2
1.0
0.8
0.6
0.4
0.2
Unknown
BCL11A
PAPOLG
REL
PUS10
Position on Chromosome 2

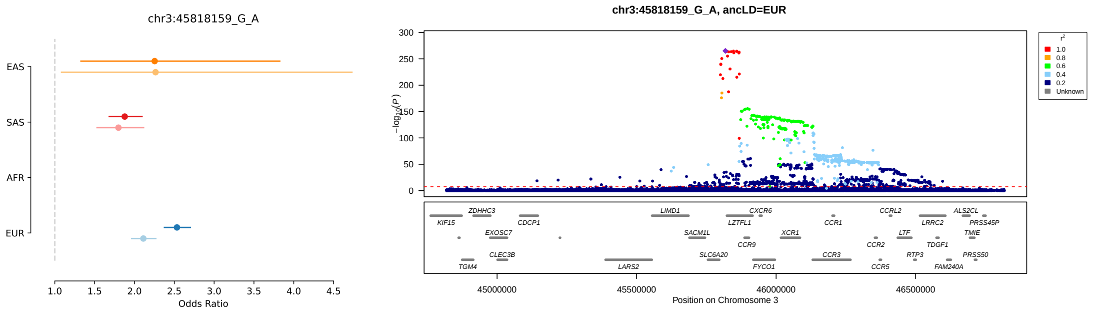
chr3:45818159_G_A
EAS
SAS
AFR
EUR
Odds Ratio
chr3:45818159_G_A, ancLD=EUR
−log10(P)
r2
1.0
0.8
0.6
0.4
0.2
Unknown
KIF15
ZDHHC3
EXOSC7
TGM4
CLEC3B
CDCP1
LARS2
LIMD1
SACM1L
SLC6A20
LZTFL1
CCR9
CXCR6
FYCO1
XCR1
CCR1
CCR3
CCR2
CCR5
CCRL2
LTF
RTP3
LRRC2
TDGF1
FAM240A
ALS2CL
TMIE
PRSS45P
PRSS50
Position on Chromosome 3

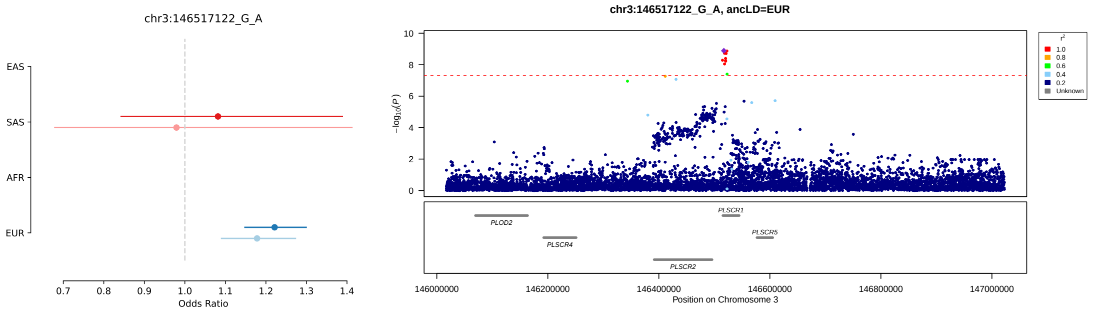
chr3:146517122_G_A
EAS
SAS
AFR
EUR
Odds Ratio
chr3:146517122_G_A, ancLD=EUR
−log10(P)
r2
1.0
0.8
0.6
0.4
0.2
Unknown
PLOD2
PLSCR4
PLSCR2
PLSCR1
PLSCR5
Position on Chromosome 3

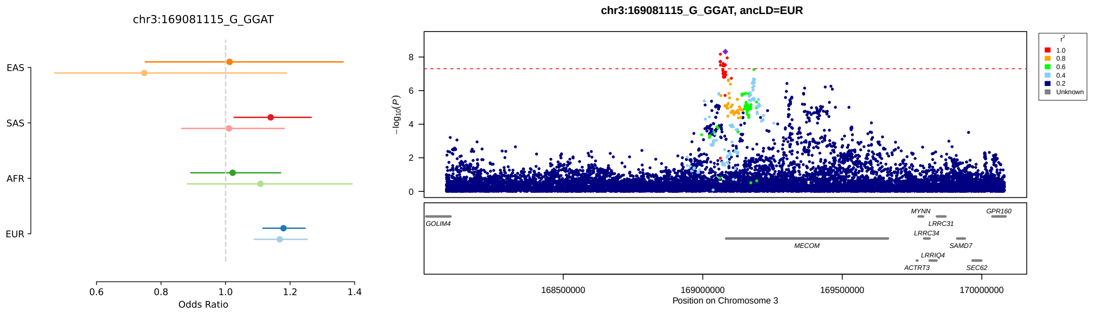
chr3:169081115_G_GGAT
EAS
SAS
AFR
EUR
Odds Ratio
chr3:169081115_G_GGAT, ancLD=EUR
−log10(P)
r2
1.0
0.8
0.6
0.4
0.2
Unknown
GOLIM4
MECOM
MYNN
LRRC34
LRRIQ4
ACTRT3
LRRC31
SAMD7
SEC62
GPR160
Position on Chromosome 3

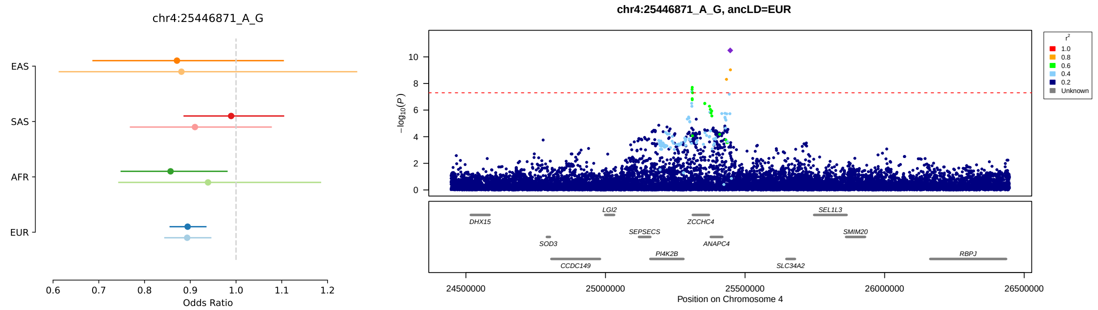
chr4:25446871_A_G
EAS
SAS
AFR
EUR
Odds Ratio
chr4:25446871_A_G, ancLD=EUR
−log10(P)
Position on Chromosome 4
r²
Unknown

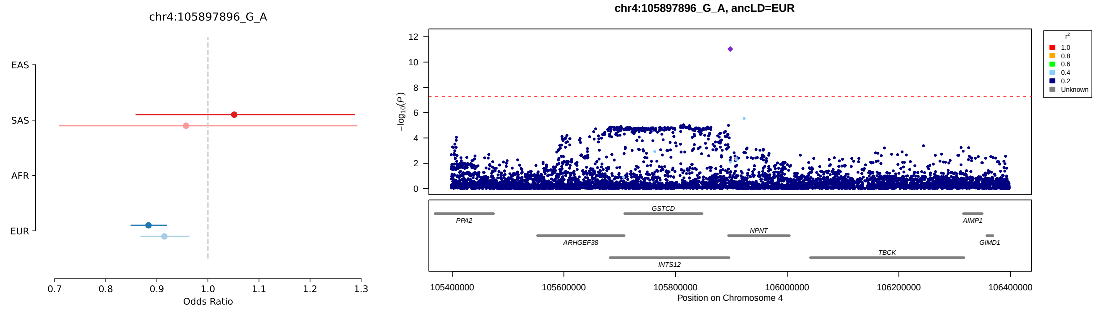
chr4:105897896_G_A
EAS
SAS
AFR
EUR
Odds Ratio
chr4:105897896_G_A, ancLD=EUR
−log10(P)
Position on Chromosome 4
r²
Unknown

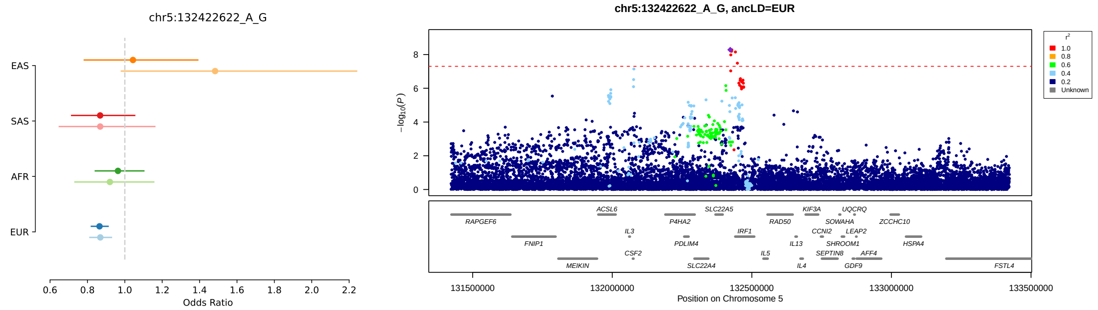
chr5:132422622_A_G
EAS
SAS
AFR
EUR
Odds Ratio
chr5:132422622_A_G, ancLD=EUR
−log10(P)
Position on Chromosome 5
r²
Unknown

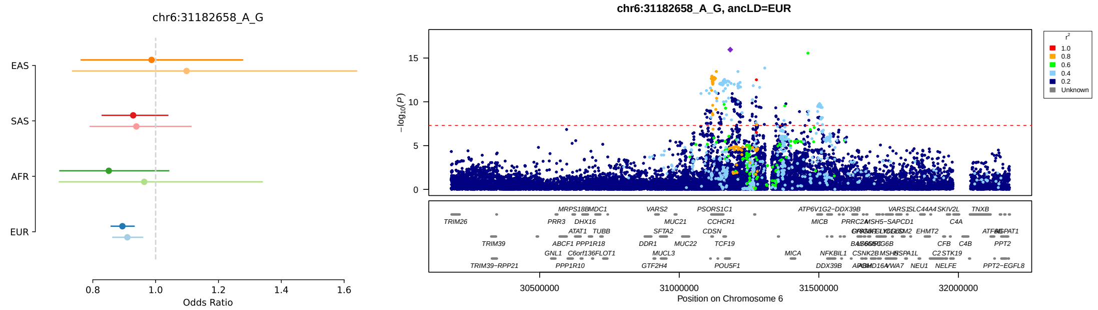
chr6:31182658_A_G
EAS
SAS
AFR
EUR
Odds Ratio
chr6:31182658_A_G, ancLD=EUR
−log10(P)
Position on Chromosome 6
r²
Unknown

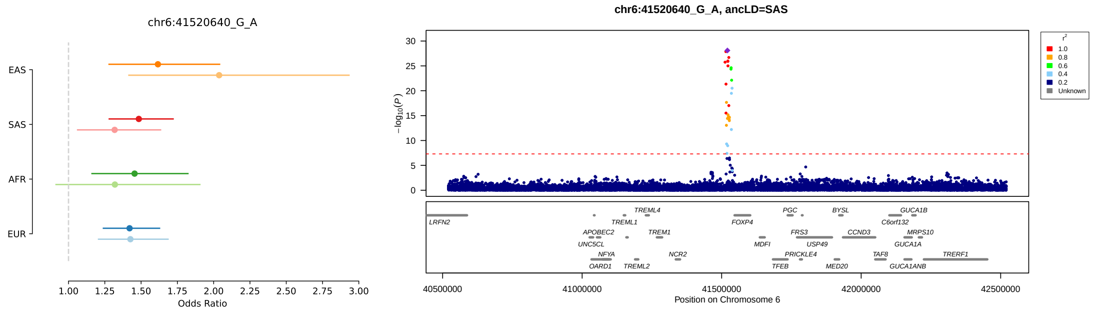

chr6:41520640_G_A
EAS
SAS
AFR
EUR
1.00 1.25 1.50 1.75 2.00 2.25 2.50 2.75 3.00
Odds Ratio
chr6:41520640_G_A, ancLD=SAS
$-\log_{10}(P)$
0 5 10 15 20 25 30
$r^2$
1.0
0.8
0.6
0.4
0.2
Unknown
LRFN2
TREML4
TREML1
APOBEC2
UNC5CL
TREM1
NFYA
OARD1
TREML2
NCR2
FOXP4
MDFI
PGC
FRS3
PRICKLE4
TFEB
BYSL
USP49
MED20
CCND3
TAF8
GUCA1B
C6orf132
MRPS10
GUCA1A
GUCA1ANB
TRERF1
40500000 41000000 41500000 42000000 42500000
Position on Chromosome 6

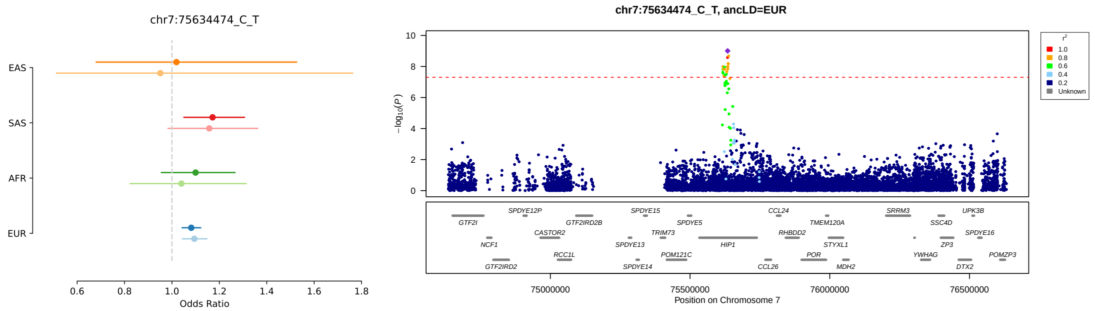

chr7:75634474_C_T
EAS
SAS
AFR
EUR
0.6 0.8 1.0 1.2 1.4 1.6 1.8
Odds Ratio
chr7:75634474_C_T, ancLD=EUR
$-\log_{10}(P)$
0 2 4 6 8 10
$r^2$
1.0
0.8
0.6
0.4
0.2
Unknown
GTF2I
NCF1
GTF2IRD2
SPDYE12P
CASTOR2
RCC1L
GTF2IRD2B
SPDYE15
SPDYE13
SPDYE14
TRIM73
POM121C
SPDYE5
HIP1
CCL24
RHBDD2
CCL26
POR
TMEM120A
STYXL1
MDH2
SRRM3
YWHAG
SSC4D
ZP3
DTX2
UPK3B
SPDYE16
POMZP3
75000000 75500000 76000000 76500000
Position on Chromosome 7

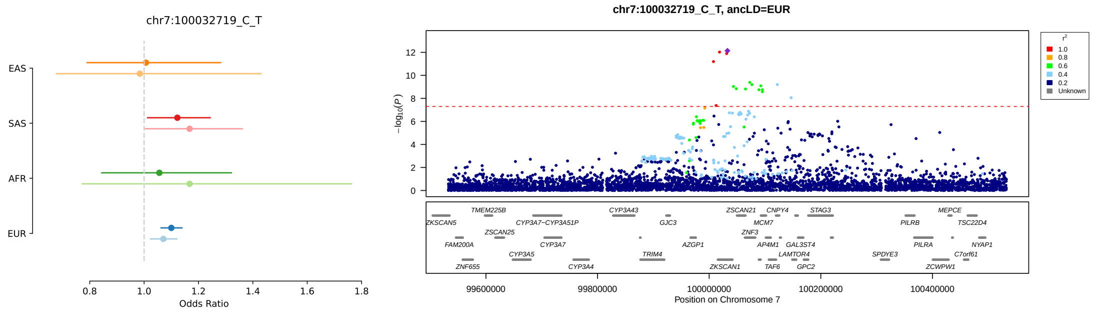

chr7:100032719_C_T
EAS
SAS
AFR
EUR
0.8 1.0 1.2 1.4 1.6 1.8
Odds Ratio
chr7:100032719_C_T, ancLD=EUR
$-\log_{10}(P)$
0 2 4 6 8 10 12
$r^2$
1.0
0.8
0.6
0.4
0.2
Unknown
ZKSCAN5
FAM200A
ZNF655
TMEM225B
ZSCAN25
CYP3A5
CYP3A7−CYP3A51P
CYP3A7
CYP3A4
CYP3A43
TRIM4
GJC3
AZGP1
ZSCAN21
MCM7
ZNF3
ZKSCAN1
CNPY4
AP4M1
TAF6
STAG3
GAL3ST4
LAMTOR4
GPC2
SPDYE3
PILRB
PILRA
ZCWPW1
MEPCE
TSC22D4
NYAP1
C7orf61
99600000 99800000 100000000 100200000 100400000
Position on Chromosome 7

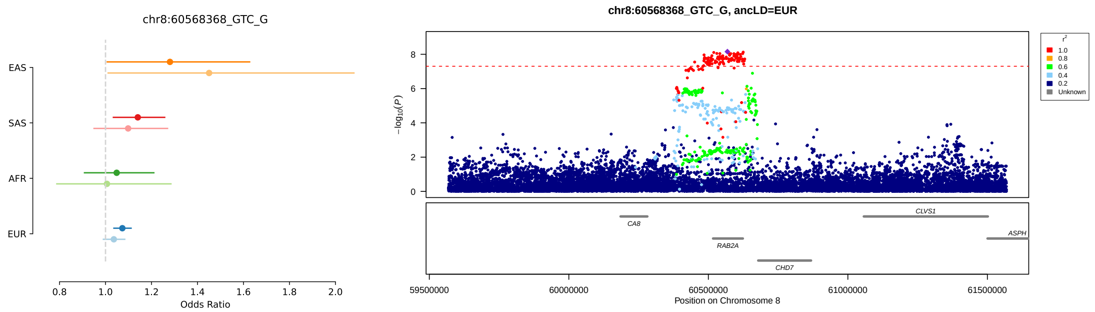

chr8:60568368_GTC_G
EAS
SAS
AFR
EUR
0.8 1.0 1.2 1.4 1.6 1.8 2.0
Odds Ratio
chr8:60568368_GTC_G, ancLD=EUR
$-\log_{10}(P)$
0 2 4 6 8
$r^2$
1.0
0.8
0.6
0.4
0.2
Unknown
CA8
RAB2A
CHD7
CLVS1
ASPH
59500000 60000000 60500000 61000000 61500000
Position on Chromosome 8

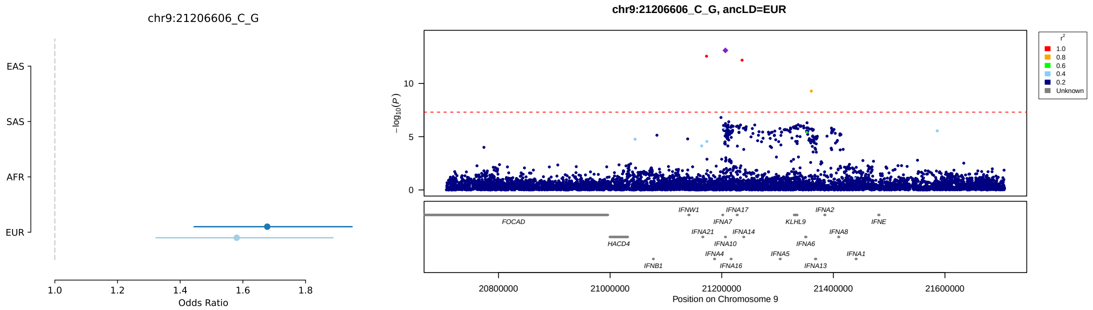
chr9:21206606_C_G
EAS
SAS
AFR
EUR
Odds Ratio
chr9:21206606_C_G, ancLD=EUR
r²
Unknown
Position on Chromosome 9

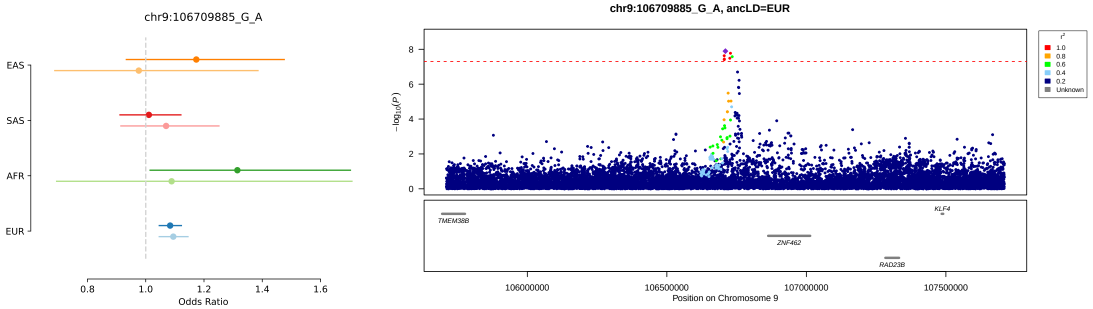
chr9:106709885_G_A
EAS
SAS
AFR
EUR
Odds Ratio
chr9:106709885_G_A, ancLD=EUR
r²
Unknown
Position on Chromosome 9

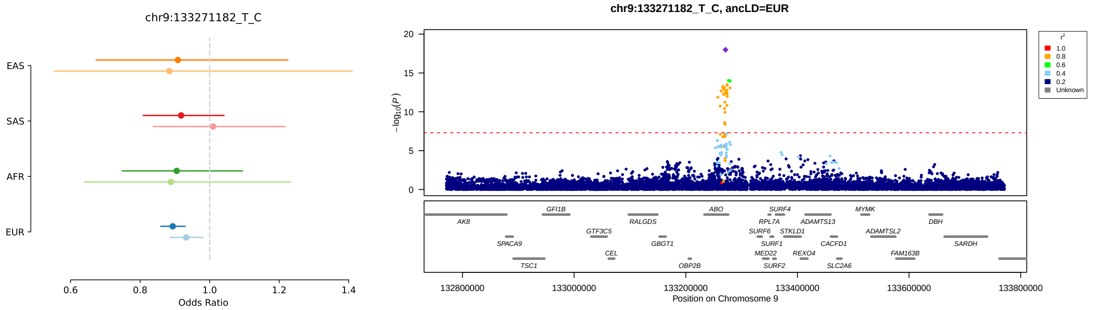
chr9:133271182_T_C
EAS
SAS
AFR
EUR
Odds Ratio
chr9:133271182_T_C, ancLD=EUR
r²
Unknown
Position on Chromosome 9

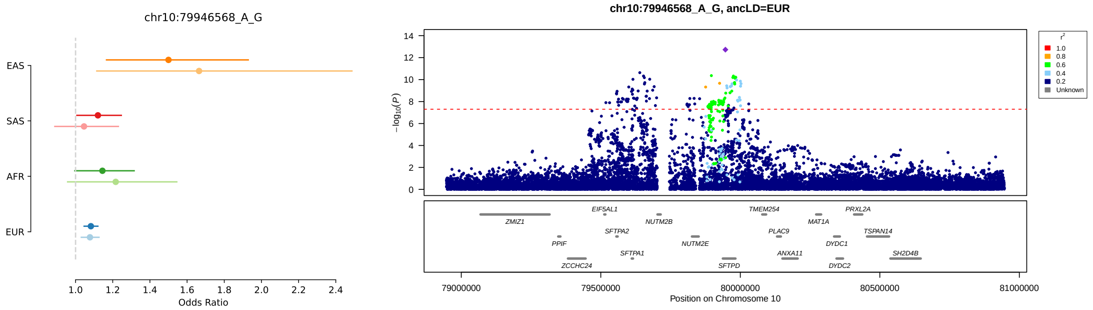
chr10:79946568_A_G
EAS
SAS
AFR
EUR
Odds Ratio
chr10:79946568_A_G, ancLD=EUR
r²
Unknown
Position on Chromosome 10

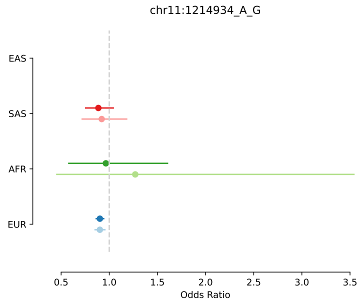
chr11:1214934_A_G
EAS
SAS
AFR
EUR
Odds Ratio
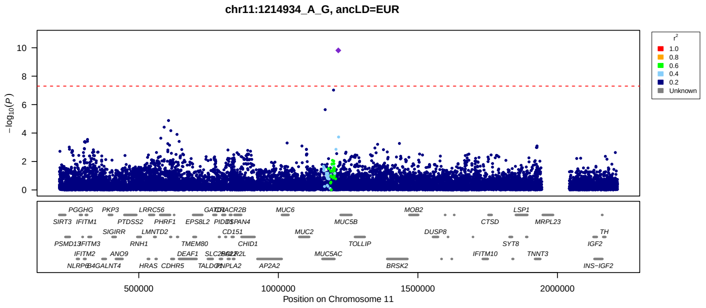
chr11:1214934_A_G, ancLD=EUR
$-\log_{10}(P)$
Position on Chromosome 11
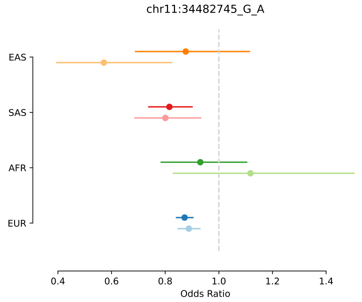
chr11:34482745_G_A
EAS
SAS
AFR
EUR
Odds Ratio
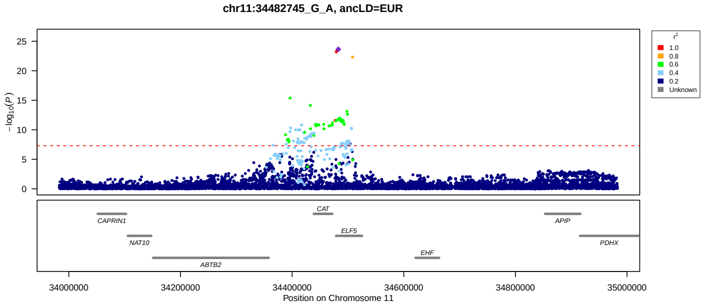
chr11:34482745_G_A, ancLD=EUR
$-\log_{10}(P)$
Position on Chromosome 11
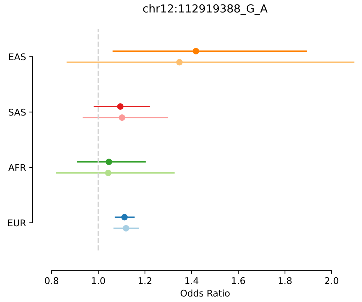
chr12:112919388_G_A
EAS
SAS
AFR
EUR
Odds Ratio
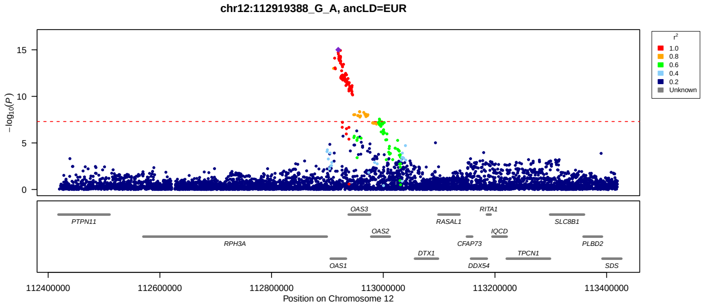
chr12:112919388_G_A, ancLD=EUR
$-\log_{10}(P)$
Position on Chromosome 12
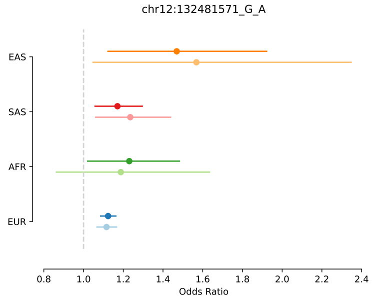
chr12:132481571_G_A
EAS
SAS
AFR
EUR
Odds Ratio
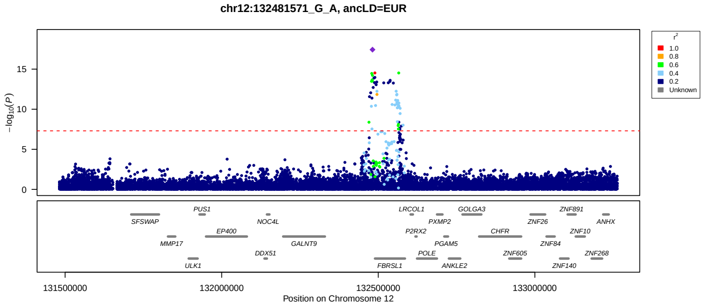
chr12:132481571_G_A, ancLD=EUR
$-\log_{10}(P)$
Position on Chromosome 12

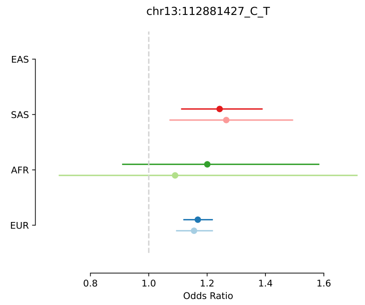
chr13:112881427_C_T
EAS
SAS
AFR
EUR
0.8
1.0
1.2
1.4
1.6
Odds Ratio

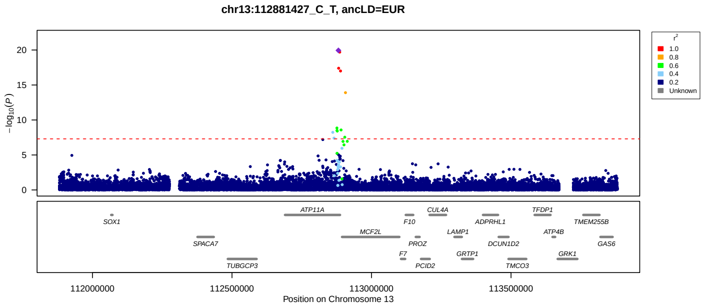
chr13:112881427_C_T, ancLD=EUR
$-\log_{10}(P)$
r²
1.0
0.8
0.6
0.4
0.2
Unknown
SOX1
SPACA7
TUBGCP3
ATP11A
MCF2L
F7
F10
PROZ
PCID2
CUL4A
LAMP1
GRTP1
ADPRHL1
DCUN1D2
TMCO3
TFDP1
ATP4B
GRK1
TMEM255B
GAS6
112000000
112500000
113000000
113500000
Position on Chromosome 13

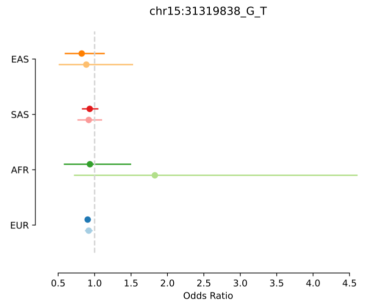
chr15:31319838_G_T
EAS
SAS
AFR
EUR
Odds Ratio

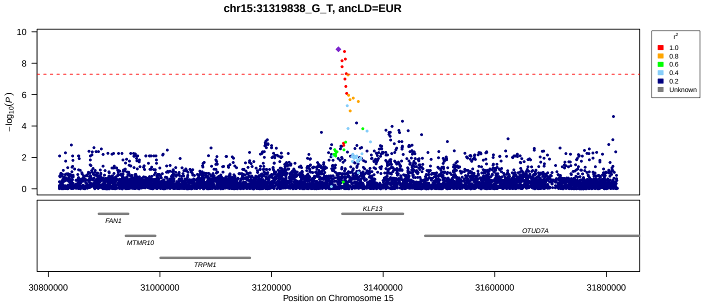
chr15:31319838_G_T, ancLD=EUR
$-\log_{10}(P)$
r²
1.0
0.8
0.6
0.4
0.2
Unknown
FAN1
MTMR10
TRPM1
KLF13
OTUD7A
30800000
31000000
31200000
31400000
31600000
31800000
Position on Chromosome 15

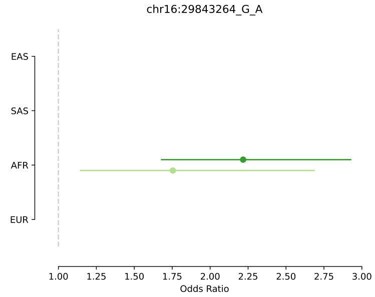
chr16:29843264_G_A
EAS
SAS
AFR
EUR
Odds Ratio

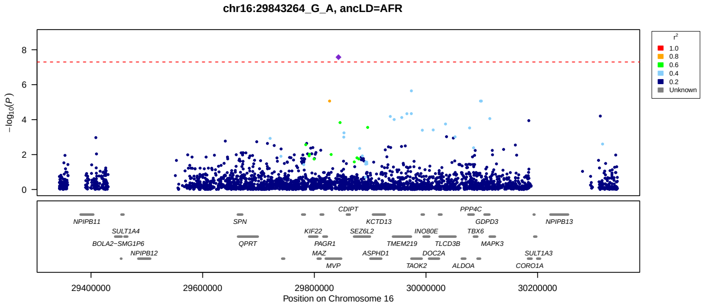
chr16:29843264_G_A, ancLD=AFR
$-\log_{10}(P)$
r²
1.0
0.8
0.6
0.4
0.2
Unknown
NPIPB11
SULT1A4
BOLA2−SMG1P6
NPIPB12
SPN
QPRT
KIF22
PAGR1
MAZ
MVP
CDIPT
SEZ6L2
ASPHD1
KCTD13
TMEM219
TAOK2
INO80E
DOC2A
TLCD3B
ALDOA
PPP4C
TBX6
GDPD3
MAPK3
NPIPB13
SULT1A3
CORO1A
29400000
29600000
29800000
30000000
30200000
Position on Chromosome 16

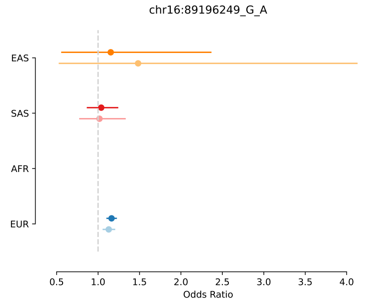
chr16:89196249_G_A
EAS
SAS
AFR
EUR
Odds Ratio

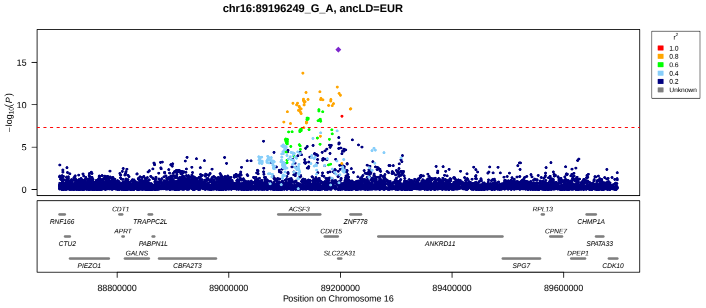
chr16:89196249_G_A, ancLD=EUR
$-\log_{10}(P)$
r²
1.0
0.8
0.6
0.4
0.2
Unknown
RNF166
CTU2
PIEZO1
CDT1
APRT
GALNS
TRAPPC2L
PABPN1L
CBFA2T3
ACSF3
CDH15
SLC22A31
ZNF778
ANKRD11
SPG7
RPL13
CPNE7
DPEP1
CHMP1A
SPATA33
CDK10
88800000
89000000
89200000
89400000
89600000
Position on Chromosome 16

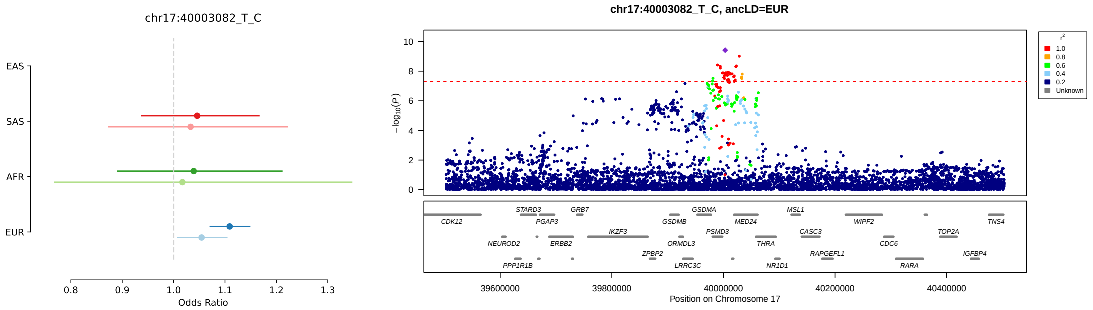
chr17:40003082_T_C
EAS
SAS
AFR
EUR
Odds Ratio
chr17:40003082_T_C, ancLD=EUR
−log10(P)
Position on Chromosome 17
r²
1.0
0.8
0.6
0.4
0.2
Unknown
CDK12
STARD3
PGAP3
GRB7
NEUROD2
ERBB2
PPP1R1B
IKZF3
ZPBP2
GSDMB
ORMDL3
LRRC3C
GSDMA
PSMD3
MED24
THRA
NR1D1
MSL1
CASC3
RAPGEFL1
WIPF2
CDC6
RARA
TOP2A
IGFBP4
TNS4

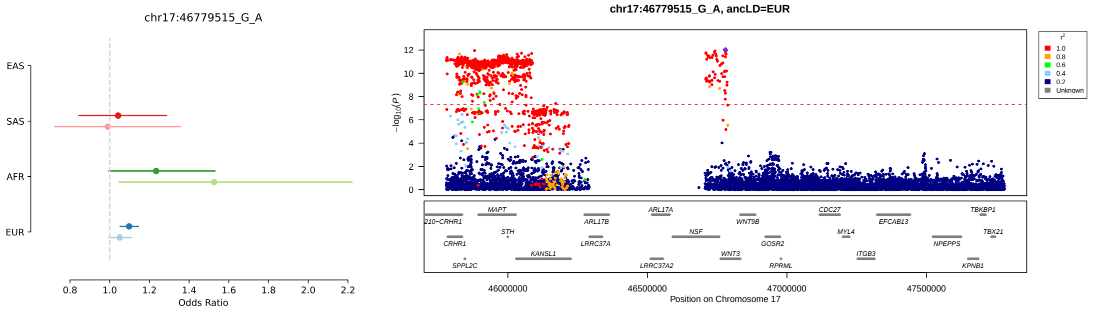
chr17:46779515_G_A
EAS
SAS
AFR
EUR
Odds Ratio
chr17:46779515_G_A, ancLD=EUR
−log10(P)
Position on Chromosome 17
r²
1.0
0.8
0.6
0.4
0.2
Unknown
MAPT
210−CRHR1
CRHR1
SPPL2C
STH
KANSL1
ARL17B
LRRC37A
ARL17A
LRRC37A2
NSF
WNT3
WNT9B
GOSR2
RPRML
CDC27
MYL4
ITGB3
EFCAB13
NPEPPS
TBKBP1
TBX21
KPNB1

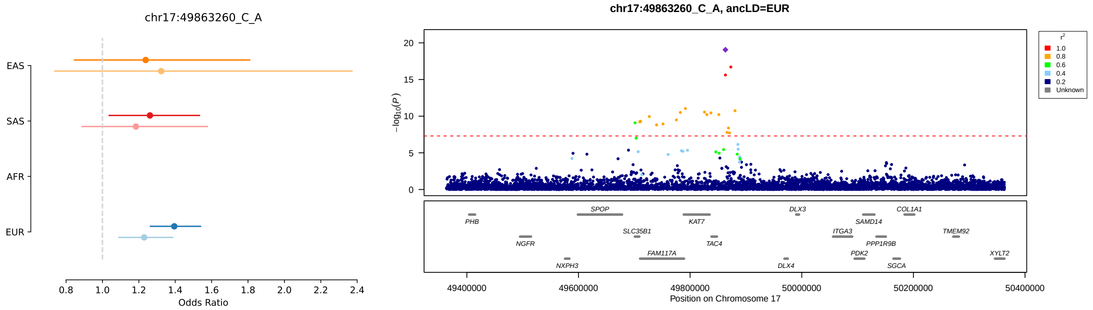
chr17:49863260_C_A
EAS
SAS
AFR
EUR
Odds Ratio
chr17:49863260_C_A, ancLD=EUR
−log10(P)
Position on Chromosome 17
r²
1.0
0.8
0.6
0.4
0.2
Unknown
PHB
NGFR
NXPH3
SPOP
SLC35B1
FAM117A
KAT7
TAC4
DLX3
DLX4
ITGA3
PDK2
SAMD14
PPP1R9B
SGCA
COL1A1
TMEM92
XYLT2

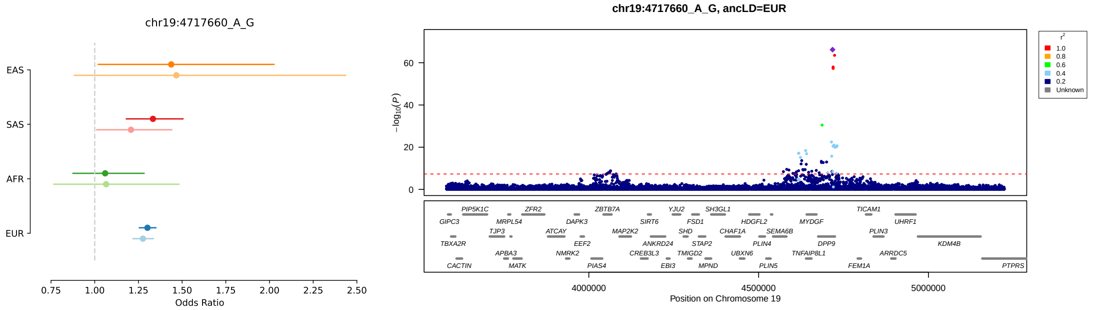
chr19:4717660_A_G
EAS
SAS
AFR
EUR
Odds Ratio
chr19:4717660_A_G, ancLD=EUR
−log10(P)
Position on Chromosome 19
r²
1.0
0.8
0.6
0.4
0.2
Unknown
GIPC3
TBXA2R
CACTIN
PIP5K1C
TJP3
APBA3
MRPL54
MATK
ZFR2
ATCAY
NMRK2
DAPK3
EEF2
PIAS4
ZBTB7A
MAP2K2
CREB3L3
SIRT6
ANKRD24
EBI3
YJU2
SHD
FSD1
TMIGD2
STAP2
MPND
SH3GL1
CHAF1A
UBXN6
HDGFL2
PLIN4
PLIN5
SEMA6B
TNFAIP8L1
MYDGF
DPP9
FEM1A
TICAM1
PLIN3
ARRDC5
UHRF1
KDM4B
PTPRS

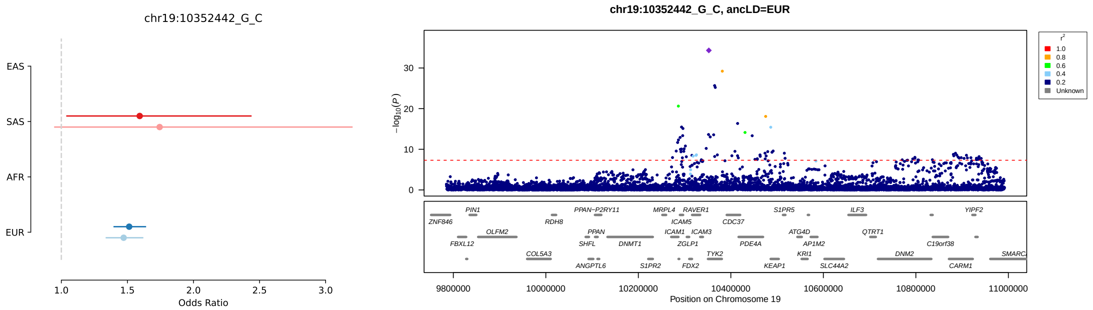
chr19:10352442_G_C
EAS
SAS
AFR
EUR
Odds Ratio
chr19:10352442_G_C, ancLD=EUR
Position on Chromosome 19

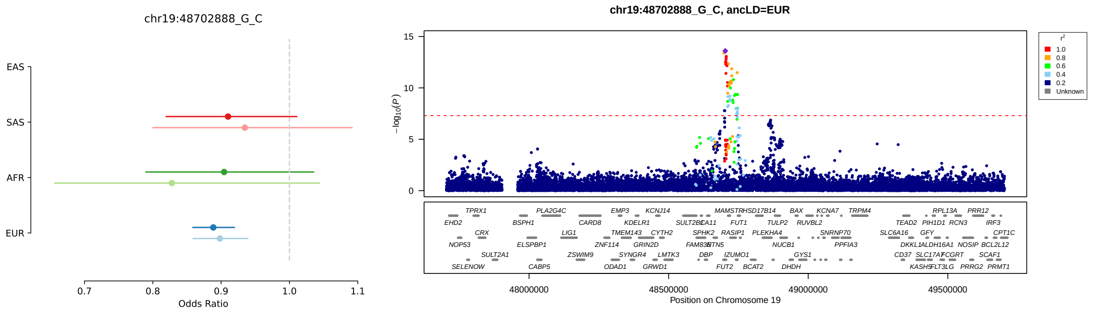
chr19:48702888_G_C
EAS
SAS
AFR
EUR
Odds Ratio
chr19:48702888_G_C, ancLD=EUR
Position on Chromosome 19

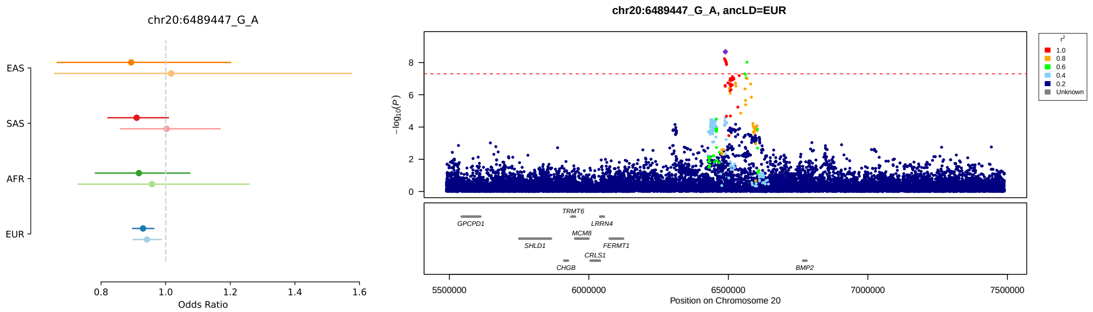
chr20:6489447_G_A
EAS
SAS
AFR
EUR
Odds Ratio
chr20:6489447_G_A, ancLD=EUR
Position on Chromosome 20

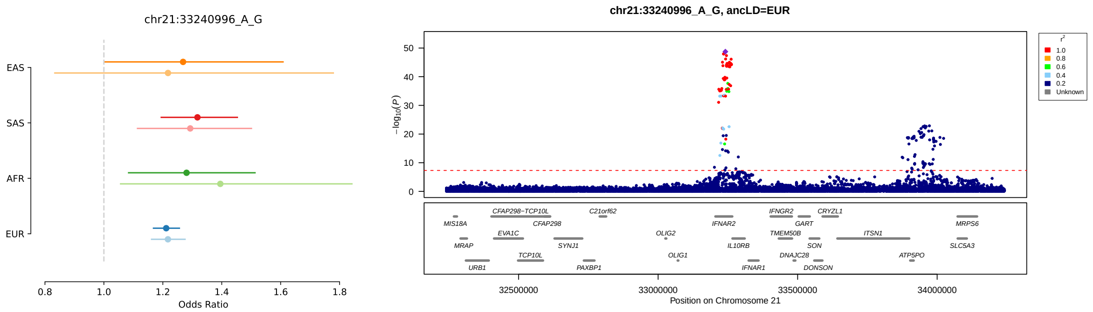
chr21:33240996_A_G
EAS
SAS
AFR
EUR
Odds Ratio
chr21:33240996_A_G, ancLD=EUR
Position on Chromosome 21

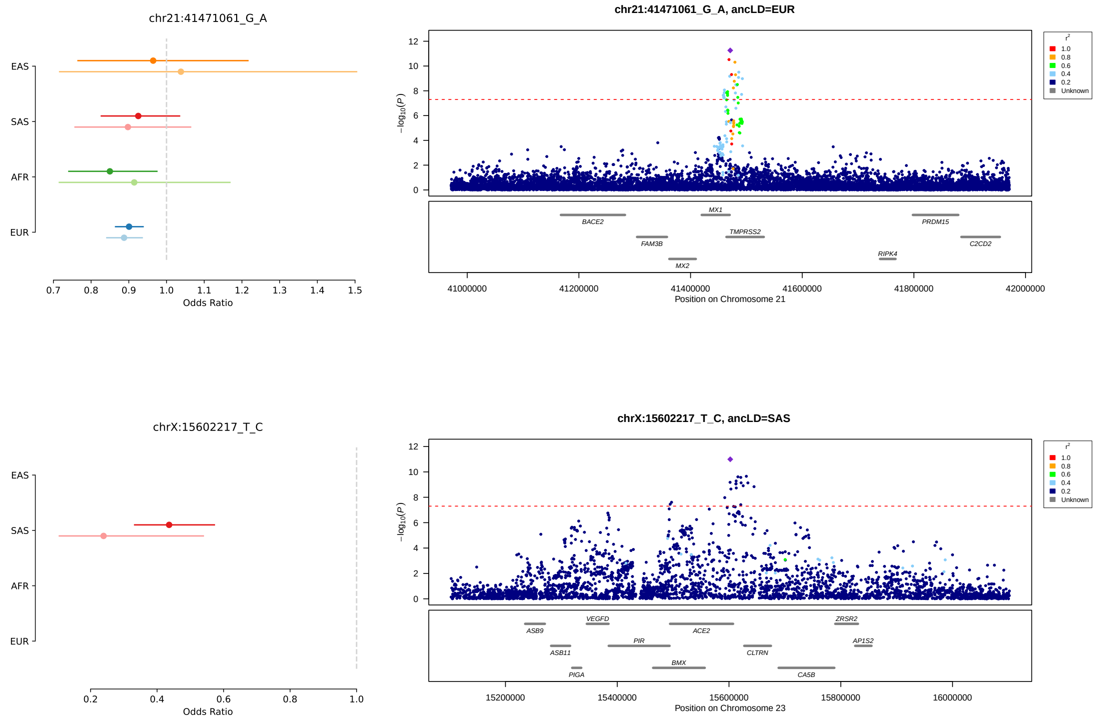

Figure 9: LocusZoom and forest plots illustrating all lead genetic association signals listed in Table 2. Panels on the right show LocusZoom plots centered on lead variant. $P$-values are from the cross-ancestry GenOMICC severe versus ctrl-all meta-analysis. The population used to calculate LD is indicated on the title and corresponds to the population having the smallest $P$-value in the GenOMICC meta-analysis. Panels on the left compare cross-ancestry odds ratios of lead variants from analyses using all controls (ctrl-all, darker lines) vs. analyses using only controls with mild COVID-19 (ctrl-mld, lighter lines).

### 3 Acknowledgments

Genomics England: This research was made possible through access to data in the National Genomic Research Library[17], which is managed by Genomics England Limited (a wholly owned company of the Department of Health and Social Care). The National Genomic Research Library (`https://www.genomicsengland.co.uk/research`) holds data provided by patients and collected by the NHS as part of their care and data collected as part of their participation in research. The National Genomic Research Library is funded by the National Institute for Health Research and NHS England. The Wellcome Trust, Cancer Research UK and the Medical Research Council have also funded research infrastructure. REACT: National Institute for Health and Care Research (NIHR) and UK Research and Innovation (UKRI) - REACT-Genomics England (REACT-GE) (MR/V030841/1) and REACT-Long COVID (REACT-LC) (COV-LT-0040). The REACT study was funded by the UK Department of Health and Social Care with supplemental funding from the Huo Family Foundation.

#### 4 Conflicts of Interest

Erola Pairo-Castineira is an employee and shareholder of Regeneron Pharmaceuticals, Inc. No Regeneron resources were used in this publication.

#### 5 Extended References

[17] National Genomic Research Library (2017).
